# A Whole Exome Sequencing Study of a small Indian Autosomal Dominant Polycystic Kidney Disease Patient Cohort

**DOI:** 10.1101/2023.04.20.23288719

**Authors:** Chandra Devi, Shivendra Singh, Bhagyalaxmi Mohapatra, Ashok Kumar, Sanjay Vikrant, Rana Gopal Singh, Pradeep Kumar Rai, Parimal Das

## Abstract

Autosomal Dominant Polycystic Kidney Disease is characterized by renal cyst development, often leading to kidney enlargement and failure. We conducted whole exome sequencing on 14 participants (12 families) from an Indian cohort. Our analysis revealed a spectrum of genetic variants, predominantly in the *PKD1.* These in *PKD1* included missense variants such as p.Glu2937Lys (c.8809G>A) and p.Gly2310Arg (c.6928G>A), p.Asp2095Gly (c.6284A>G), p.Thr938Met (c.2813C>T), p.Trp967Arg (c.2899T>C), p.Glu593* (c.1777G>T), frameshift variants p.Gln149fs*141 (c.445delC), p.Ser3305fs*84 (c.9914_9915delCT), p.His1347fs*83 (c.4041_4042delCA), and p.Leu2776fs*87(c.8327_8363delTGGCGGGCGAGGAGATCGTGGCCCAGGGCAAGCGCTC), intronic splice site variant c.8017-3C>G, nonsense variant p.Glu593* (c.1777G>T) and in *PKD2* missense variant p.Ser370Asn (c.1109G>A). While one individual carried intronic (c.2358+5G>A) and 3’UTR (c.*174G>T) variants in *PKD2* only another individual carried variants in both *PKD1* and *PKD2*, suggesting potential genetic complexity. Clinical data revealed diverse presentations. Age at diagnosis varied widely. Patients with frameshift variants exhibited earlier onset and severe manifestations, including bilateral ADPKD. One proband had right unilateral ADPKD. Involvement of liver, a common extra-renal manifestation, was also observed. Heterogeneity at phenotypic and at allelic level was observed in our cohort. In this study, using WES of a trio, a frameshift-truncation deletion [c.32del/p.Leu11ArgfsTer61] in *MIOX* was found to be associated with the disease shared by both the affected and early diagnosed mother and daughter carrying *PKD1* missense variant, which had not been previously reported in ADPKD. Further, differential gene expression analysis using data from GEO database showed reduced MIOX expression in ADPKD cystic samples compared to minimal cystic tissues and controls. MIOX is an enzyme specific to renal tubules and catalyses the initial step of the kidney-based myoinositol catabolism. Both affected candidates also shared benign variants and other variations of uncertain significance which may influence the disease development. Further functional analysis will clarify how MIOX contributes to the disease. The study limitations include the small sample size and the need for validation in larger cohorts. Our findings highlight the importance of genetic analysis in ADPKD management especially to facilitate personalized therapeutic strategies.

**Highlights:**

- Identified variants in *PKD1* and *PKD2* through whole exome sequencing in ADPKD patients, affecting different protein regions.
- Variants include non-synonymous coding changes, frame-shift deletions, and splice site alterations.
- Clinical features and age at diagnosis varied widely, with common symptoms including flank pain, fatigue.
- Frameshift deletion in *MIOX*, associated in one PKD1 trio, implicates its role in ADPKD pathogenesis.
- DGE analysis of dataset from database reveals downregulation of MIOX in ADPKD tissue samples highlighting its role in potential molecular pathways in ADPKD progression.

**Graphical abstract:** 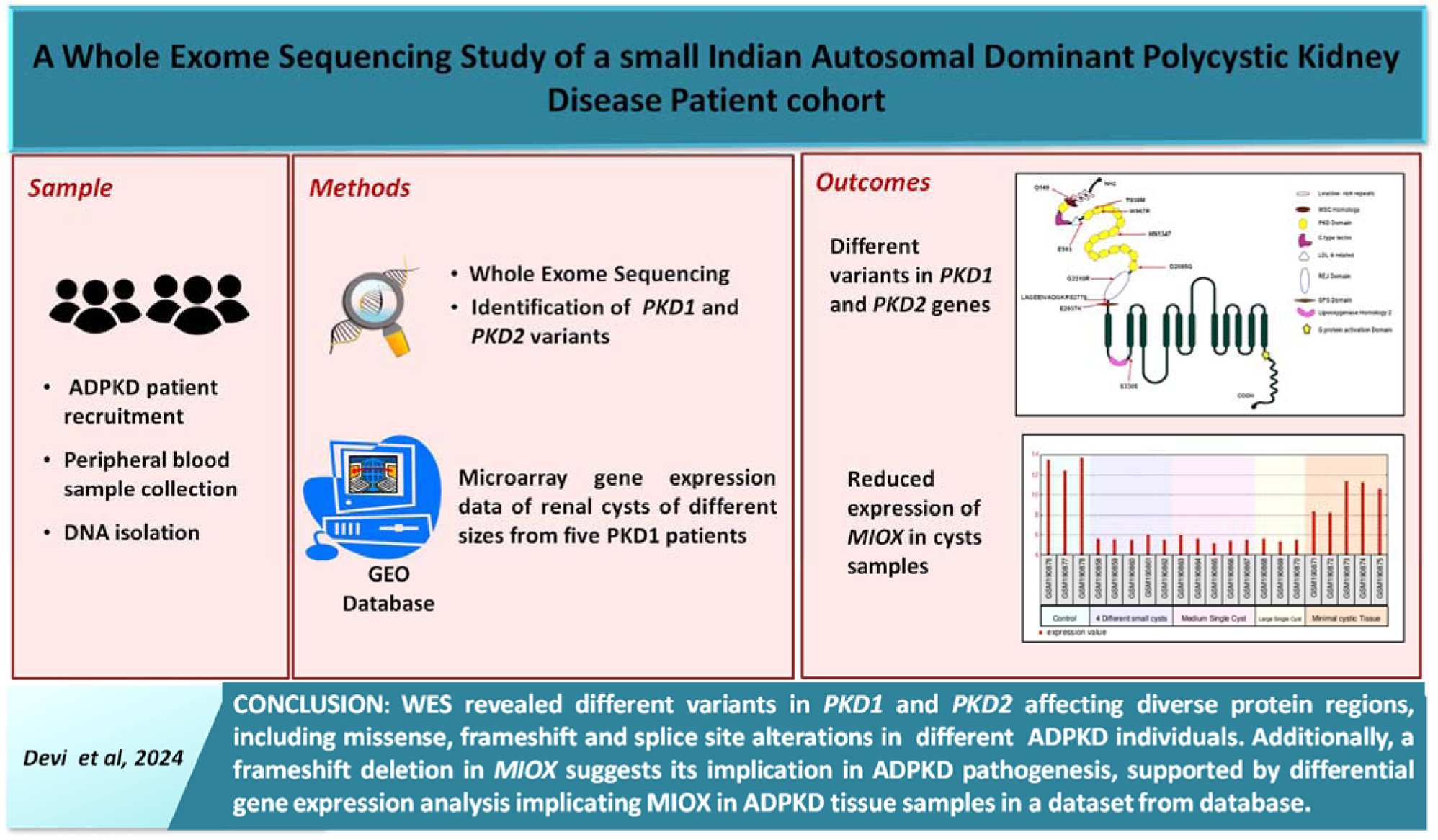

## Introduction

Autosomal Dominant Polycystic Kidney Disease (ADPKD) is a common hereditary renal disorder affecting 1 in 500 - 1,000 individuals worldwide and is characterized by the development of multiple fluid-filled cysts in the kidneys, leading to progressive renal dysfunction [1, 2]. ADPKD is primarily caused by mutations in the *PKD1* (∼75%) and *PKD2* (∼15% −18%) which encode for polycystin-1 (PC1) and polycystin-2 (PC2) proteins, respectively and the remaining 7-10% of the cases remain genetically unresolved, and sporadic and mosaic ADPKD cases have also been observed [3–5]. This condition is genetically and phenotypically diverse, comprising a wide range of signs and severity levels even within the same family [6]. The prevalence of PKD also varies among different ethnic groups. Key features include renal cysts leading to kidney enlargement and failure, in many cases liver cysts causing hepatomegaly, and intracranial aneurysms, posing a risk of hemorrhagic stroke. In addition to these cardinal manifestations, ADPKD can also present with various extra renal manifestations and complications such as hypertension, urinary tract infections, renal stones, hematuria, proteinuria, renal insufficiency, and chronic pain. ADPKD has also been associated with an increased risk of cardiovascular abnormalities, such as left ventricular hypertrophy and valvular abnormalities, as well as abnormalities in other organs, including the pancreas, spleen, and reproductive system [7].

The diagnosis of ADPKD involves clinical evaluation and various imaging techniques, including ultrasonography, CT scans and MRI to detect renal cysts and assess their size and number [8, 9]. Genetic testing is essential, especially in cases with atypical presentations or uncertain diagnoses, to confirm pathogenic variations in the *PKD1* or *PKD2* genes and differentiate ADPKD from other kidney diseases. *PKD1* is a large gene that spans a 52kb region of genomic DNA and presents a challenge for direct PCR and Sanger sequencing for mutation analysis due to the presence of six pseudogenes [10]. Adding to the genetic complexity of the disease, recent studies have identified *GANAB*, *DNAJB11*, *ALG9*, *IFT140* as minor/associated genes [3, 5, 11]. Next-Generation Sequencing (NGS), such as Whole Exome Sequencing (WES), offers a comprehensive approach to analyzing genetic profiles, aiding in better understanding and management of PKD [12, 13]. The considerable genetic heterogeneity observed in *PKD1* variants contributes to diverse clinical presentations and disease progression among affected individuals [5, 11]. While some variants result in severe disease manifestations early in life, others exhibit a more indolent course with later onset of symptoms [14, 15]. While extensive genetic analyses have been conducted on ADPKD cohorts globally, there remains a need for further exploration, particularly in diverse populations such as the Indian population, to elucidate the full spectrum of genetic variations contributing to the disease. Accurate classification, treatment, and monitoring of the disease rely on the detection of pathogenic variants in candidate genes such as *PKD1* and *PKD2*.

In our study, we used WES to identify pathogenic variants related to ADPKD major candidate genes highlighting the importance of molecular genetics in diagnosing and managing the disease. Also, using WES of a trio, a frameshift-truncation deletion [c.32del/p.Leu11ArgfsTer61] in *MIOX* was found to be associated with the disease shared by both the early diagnosed affected mother and daughter carrying *PKD1* missense variant, which has not been previously reported in ADPKD or other polycystic kidney diseases. In addition, the differential gene expression analysis the cystic tissue samples (GEO dataset: GSE7869) revealed a remarkable reduction in *MIOX* gene expression. *MIOX* encodes Myo-Inositol Oxygenase, which expresses exclusively in renal tubules and catalyzes the first step of myoinositol catabolism pathway predominantly occurring in the kidney [16]. While further functional evaluation is needed to elucidate the precise role of this variation in disease progression, the study highlights the potential of WES as a tool for PKD diagnosis and towards enhancing the scope for practicing personalized medicine.

## Material and methods

### Sample collection and DNA isolation

The 14 participants (from 12 families) were recruited from Sir Sunderlal Hospital, Banaras Hindu University, with informed consent obtained from all participants. Each sample was assigned a unique study code to ensure confidentiality and anonymity throughout the research process. The study received approval from the Institute Ethical Committee of the Institute of Science, Banaras Hindu University. Genomic DNA was extracted from peripheral blood samples using the standardized salting-out method [17]. Assessment of DNA quality and quantity was performed using spectrophotometric analysis (Nanodrop 2000, Thermo Scientific Inc.) or fluorometry-based methods (DNA Assay BR, Invitrogen, Cat# Q32853 / Qubit High Sensitivity Assay, Invitrogen).

### Whole Exome Sequencing

WES was performed using Illumina Next-Generation Sequencing. The WES platform targeted approximately 30Mb of the human exome, covering around 99% of the regions in CCDS and RefSeq. The mean sequencing depth was 80-100X, with more than 90% coverage of the target regions at a depth of 20X. Duplicate reads were removed, and base quality was recalibrated to ensure high-quality data. The reads were aligned to the human reference genome GRCh37 (hg19). Variant identification followed the GATK best practice framework, and variants were annotated using databases such as OMIM, GWAS, GNOMAD, and 1000Genome. The annotated variants underwent filtration based on several criteria including gnomAD genome allele frequency of ≤ 0.01 or NA (not available), read depth of ≥ 20, pathogenicity, and functional impact.

### Gene Expression Data Source for Differential Gene Expression Analysis

The GEO database, GSE7869, from which the microarray gene expression data from renal cysts of different sizes of five PKD1 patients was retrieved, includes small cysts (<1 ml, n = 5), medium cysts (10–20 ml, n = 5), large cysts (>50 ml, n = 3), minimally cystic tissue (MCT, n = 5), and non-cancerous renal cortical tissue from three nephrectomized kidneys with isolated renal cell carcinoma as normal control (n = 3). The samples were examined using the Human Genome U133 Plus 2.0 Array chip platform from Affymetrix.

### Differential Gene Expression Analysis

The raw data were normalized using the using R - RMA normalization followed by BiocManager::install (version = “3.12”) BiocManager::install (“GEOquery”) BiocManager::install (“affy”) BiocManager::install (“hgu133a.db”, type = “source”) BiocManager::install (“hgu133acdf”) Calling Libraries: library (GEOquery) library (affy) library (hgu133a.db) library (hgu133acdf). The limma function package of the R language (version 3.5.2) was used for the differentially expressed gene analysis [18], and the absolute logarithmic conversion of the differential expression multiple (Log2FC) value >2 and FDR 0.05 were used as screening criteria.

## Results

### Clinical Features

Our study involved WES of 14 participants (12 probands, 1 affected relative, and 1 normal) from the Indian population, revealing different variants within the *PKD1* and *PKD2* genes. Both male and female patients were included, with ages ranging from 20 to 56 years at the time of sampling. The clinical profiles of the ADPKD patients in our cohort exhibited a spectrum of manifestations characteristic of the disease. A significant proportion of patients reported a positive family history of ADPKD, consistent with the autosomal dominant inheritance pattern of the disease. The enlarged kidney size with multiple cysts and bilateral involvement was predominant among the patients. Only one proband (individual with given ID 10_CD103) presented with unilateral kidney cysts and the sibling of this proband also exhibited unilateral kidney cysts at the teenage. Liver cysts were also noted in three cases. Patients reported a range of systemic symptoms such as flank pain, weakness, fatigue, and hematuria associated with ADPKD. These included flank pain which was a common complaint among the cohort, often attributed to cyst enlargement or hemorrhage within the kidneys. Generalized weakness and fatigue were also prevalent, reflecting the systemic impact of chronic kidney disease on overall health and vitality.

### *PKD1* Variants

By applying a stringent filter based on gnomAD genome allele frequency of ≤ 0.01 or NA (not available), we observed that the majority of ADPKD individuals (11 out of 12 probands) exhibited heterozygous variants in *PKD1* except one carrying a homozygous missense variant, signifying its predominant role in ADPKD pathogenesis in our cohort. Among these, two individuals carried the same variant in the intronic splice site region of the *PKD1* gene, while another individual carried variants in both *PKD1* and *PKD2*. One individual was found to carry a variant in the *PKD2* gene. Different variants (Table 2) in different ADPKD individuals were identified in the *PKD1* gene viz. missense variants p.Glu2937Lys (c.8809G>A) in exon 24, p.Gly2310Arg (c.6928G>A) in exon 16, synonymous variant p.Thr2710= (c.8130C>T) in exon 22, Intronic-Splice Site Region variant c.8017-3C>G in exon 21 in two individuals, nonsense variant c.1777G>T (p.Glu593*) in exon 9, frameshift variants: p.Gln149fs*141 (c.445delC) in exon 4, p.Ser3305fs*84 (c.9914_9915delCT) in exon 29, p.His1347fs*83 (c.4041_4042delCA) in exon 15, novel Codon Deletion p.Ser2792_Leu2793del (c.8374_8379delAGCCTG) in exon 23, novel p.Asp2789_Pro2790delinsGlu (c.8367_8369delCCC) Codon Change plus Codon Deletion in exon 23, novel p.Leu2776fs*87 (c.8327_8363delTGGCGGGCGAGGAGATCGTGGCCCAGGGCAAGCGCTC) Frameshift in exon 23. The frameshift variants p.Gln149fs141, p.Ser3305fs84, and p.His1347fs*83 were classified as pathogenic variants according to ACMG criteria and were not reported in population databases (Table 3).

**Table 1:**
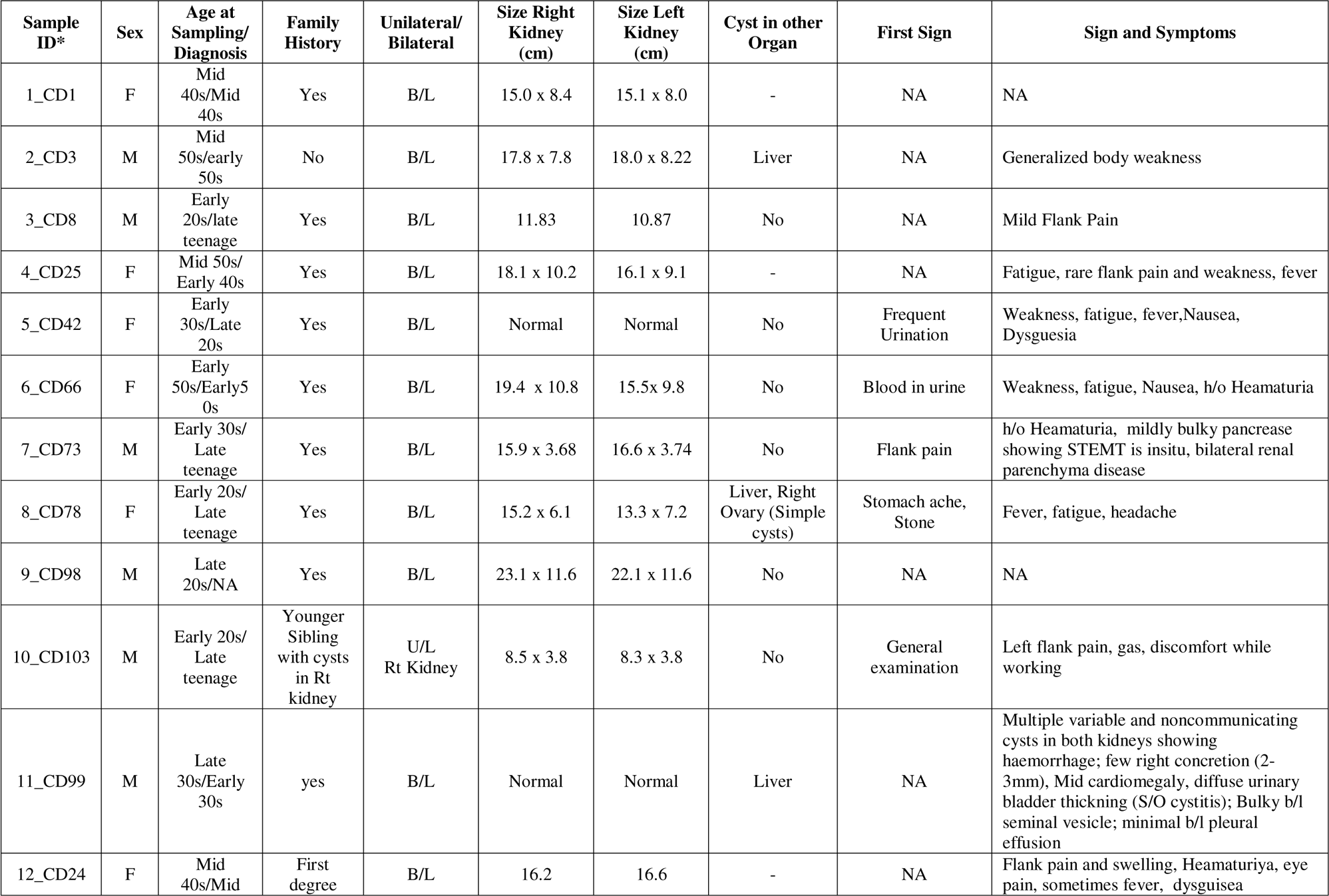
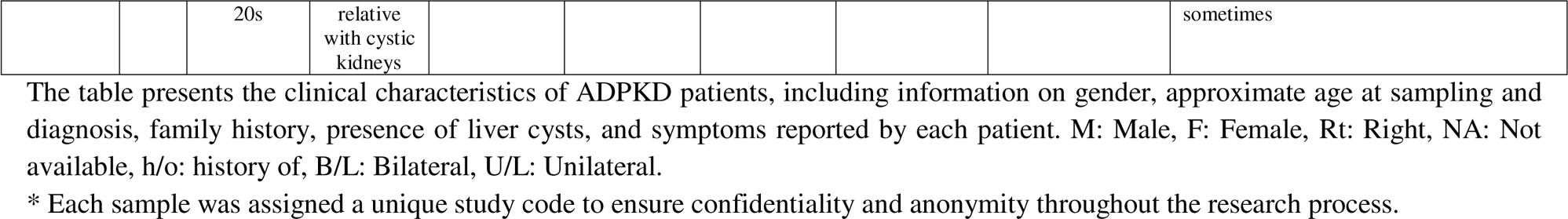
Clinical Features of Autosomal Dominant Polycystic Kidney Disease Patients.

**Table 2:**
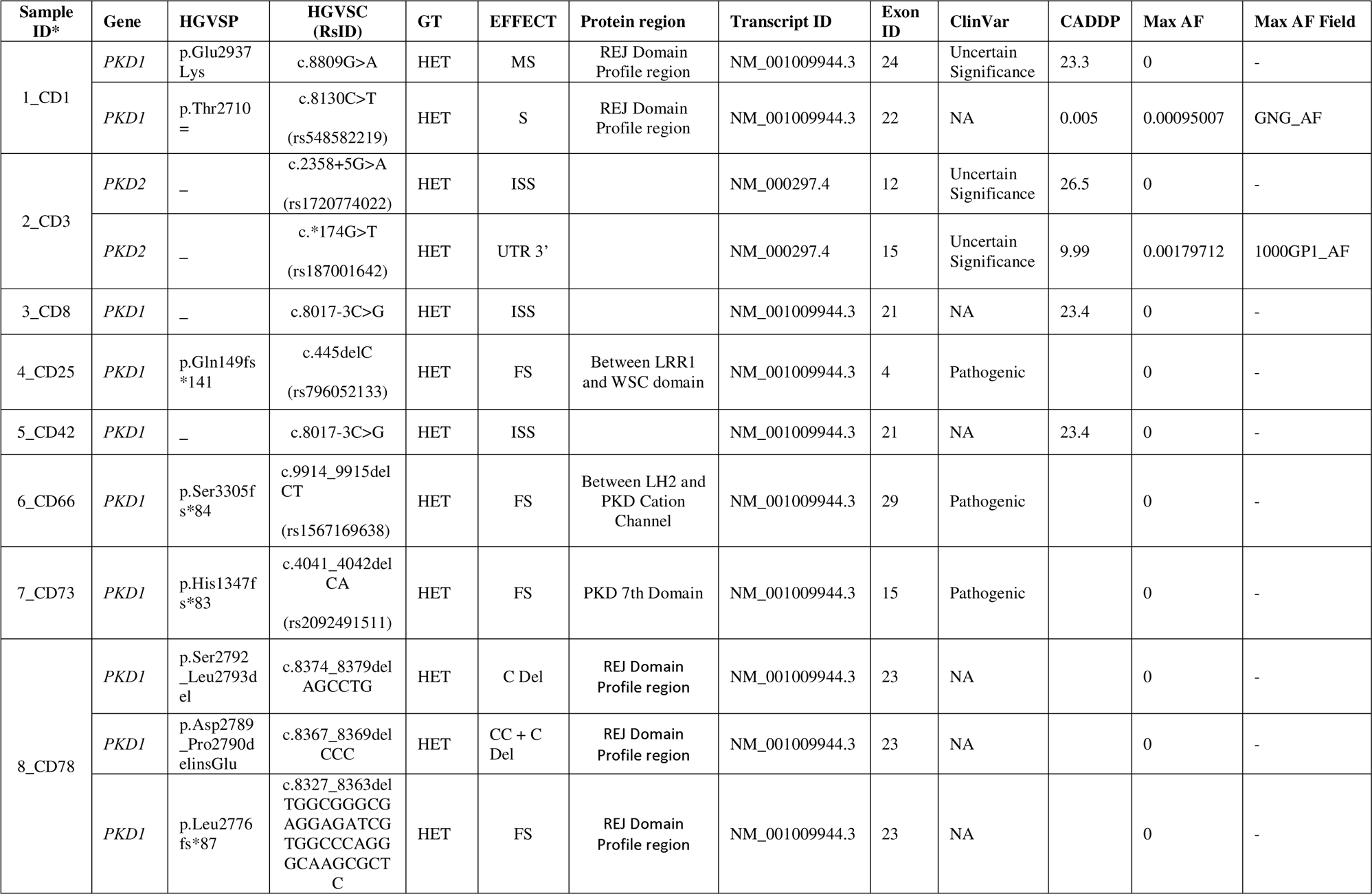
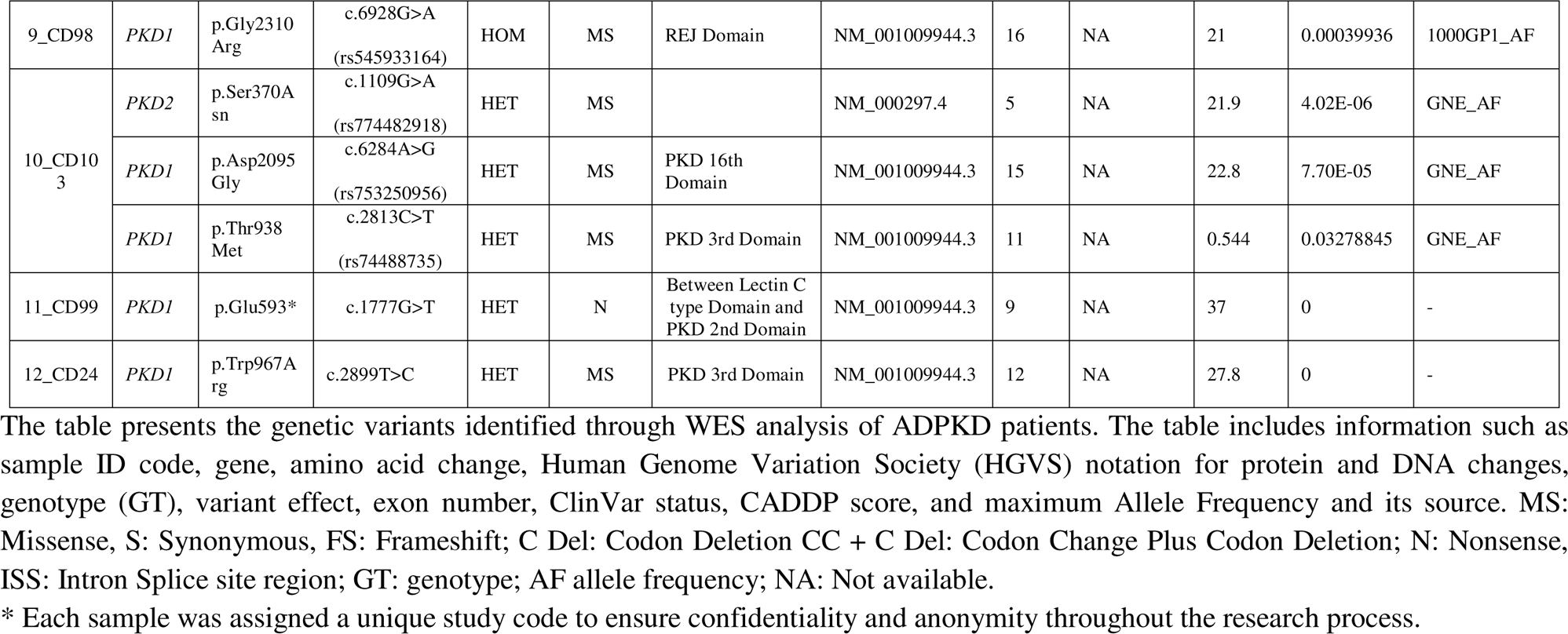
WES data showing *PKD1* and *PKD2* genetic variants identified in ADPKD patients.

**Table 3:**
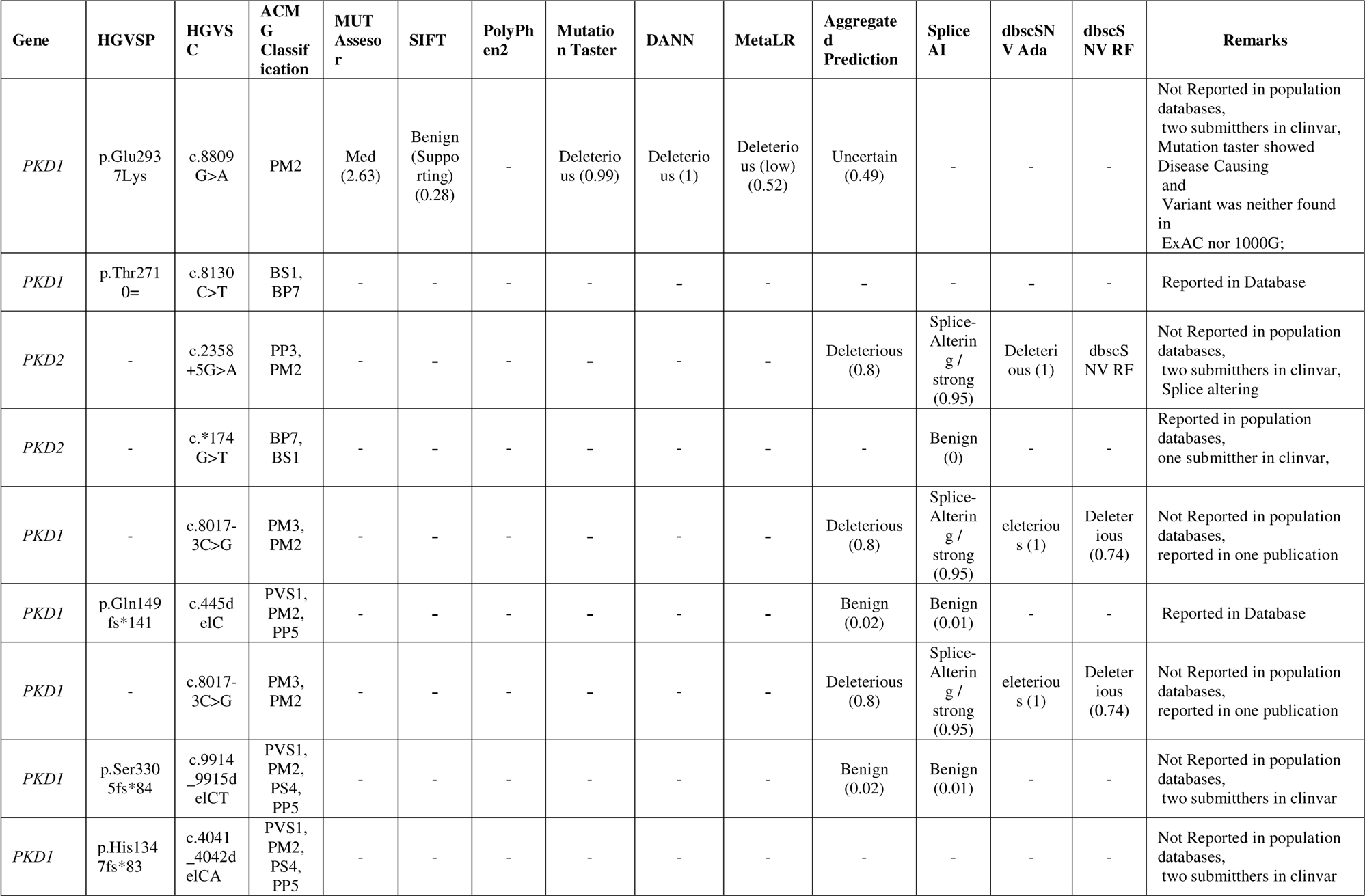
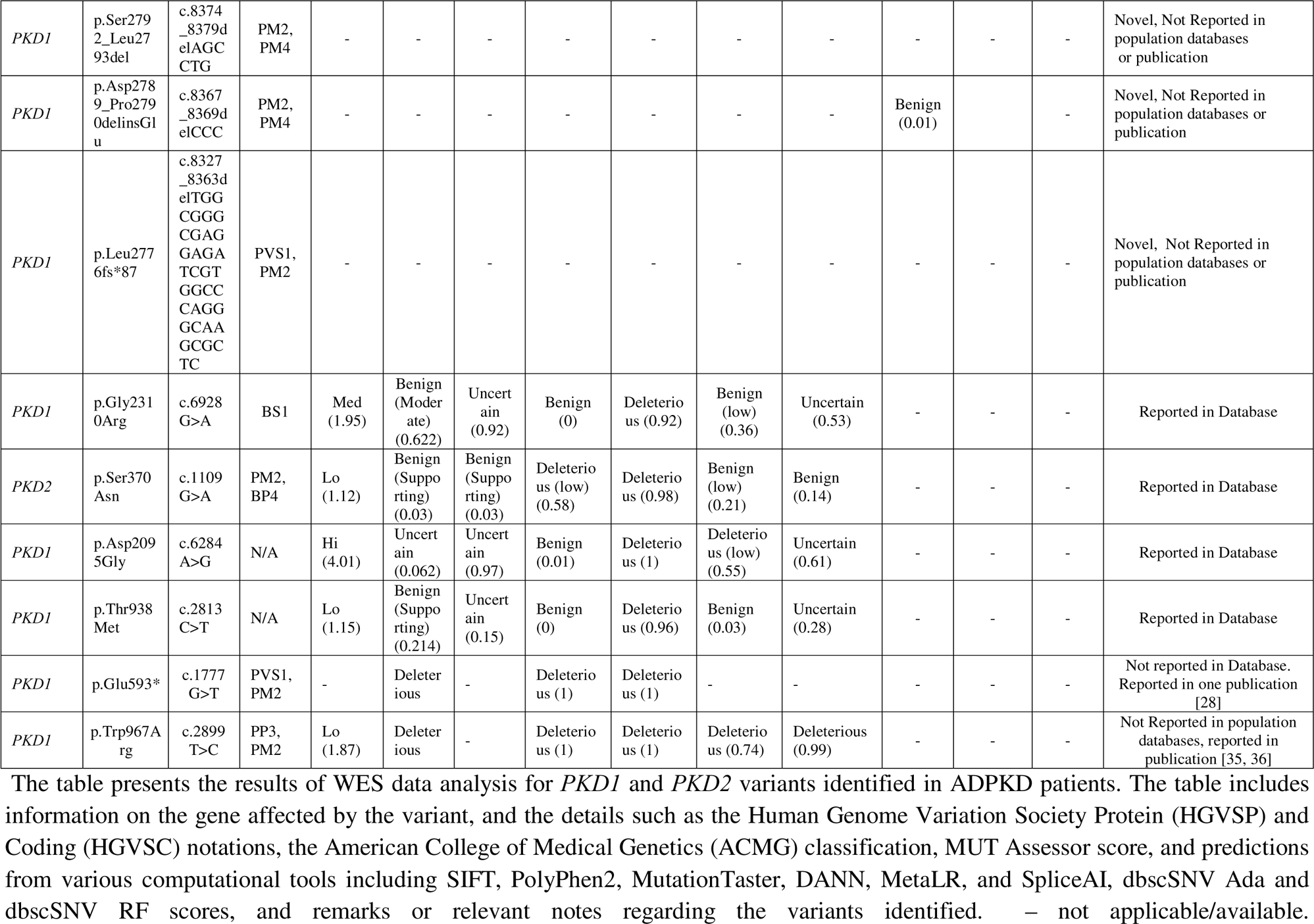
*PKD1* and *PKD2* Variants Analysis.

The exons and corresponding amino acid intervals are mentioned in table 4. The amino acid number and corresponding motif/domains are mentioned in table 5 and 6, the variants affecting the domain in protein structure are depicted in figure 1 and the anticipated functions of each domain in table7.

**Figure 1:**
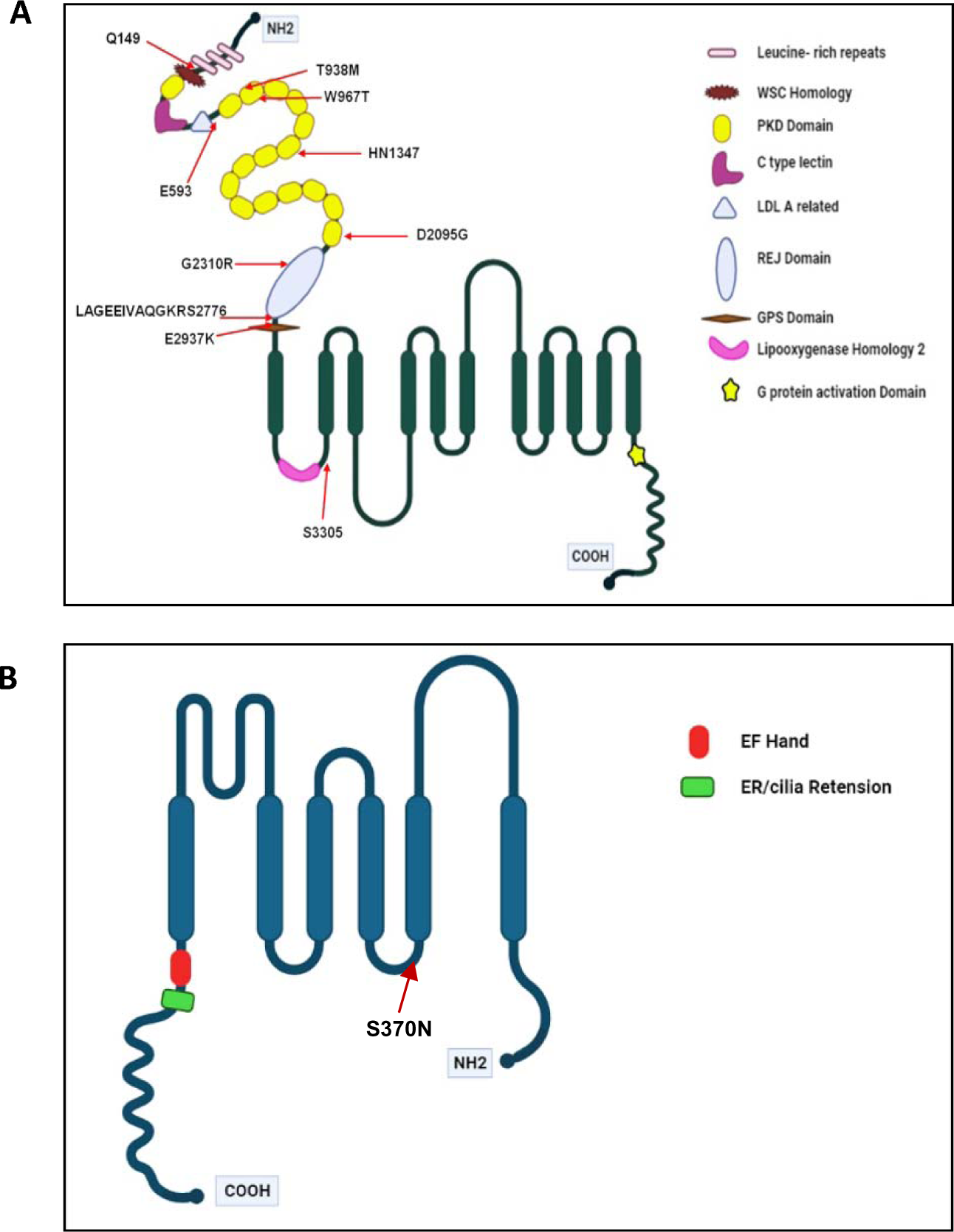
Schematic representation of the coding region variants’ location in protein structure of PKD1 and PKD2. A) PKD1/PC1 is a large transmembrane protein with multiple domains, including N-terminal region (NH_2_), leucine-rich repeat (LRR) domain, cell wall integrity and stress component (WSC) domain, C-type lectin domain, low density lipoprotein-A (LDL-A) domain, Ig-like 16 PKD repeats, receptor for egg jelly (REJ), GPCR proteolytic site (GPS) comprising N-terminal region followed by PLAT (Polycystin-1, Lipoxygenase, Alpha-Toxin) domain, 11 transmembrane domains, G protein activation domain and C-terminal coiled-coil tail. B) PKD2/PC2 is a smaller protein with six transmembrane domains (TM1-TM6) and cytoplasmic N- and C-termini. Both proteins are essential components of the polycystin complex involved in calcium signaling and regulation of renal tubular cell proliferation and differentiation.

**Table 4:**
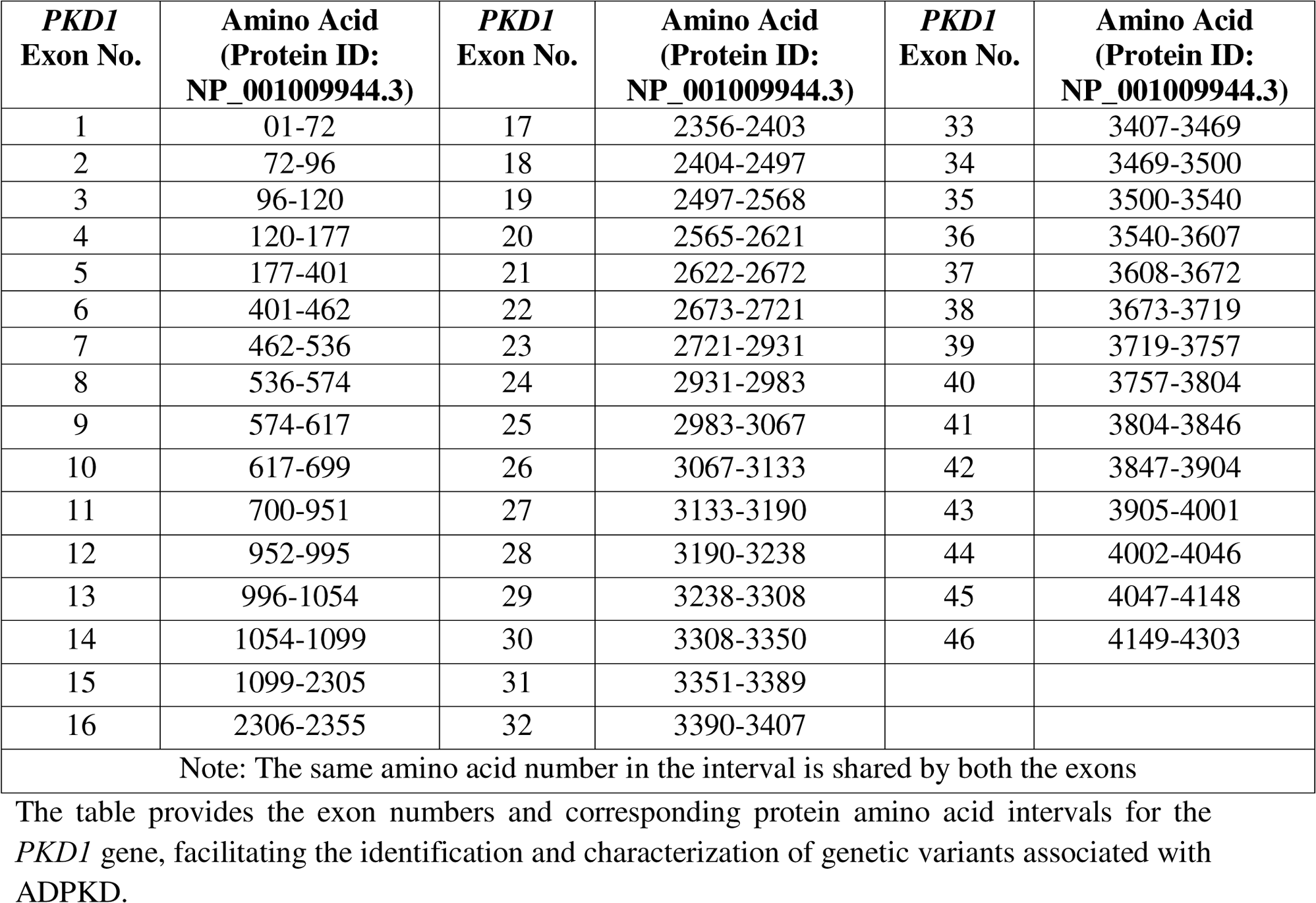
*PKD1* Exon Number and Protein Amino Acid Intervals.

**Table 5:**
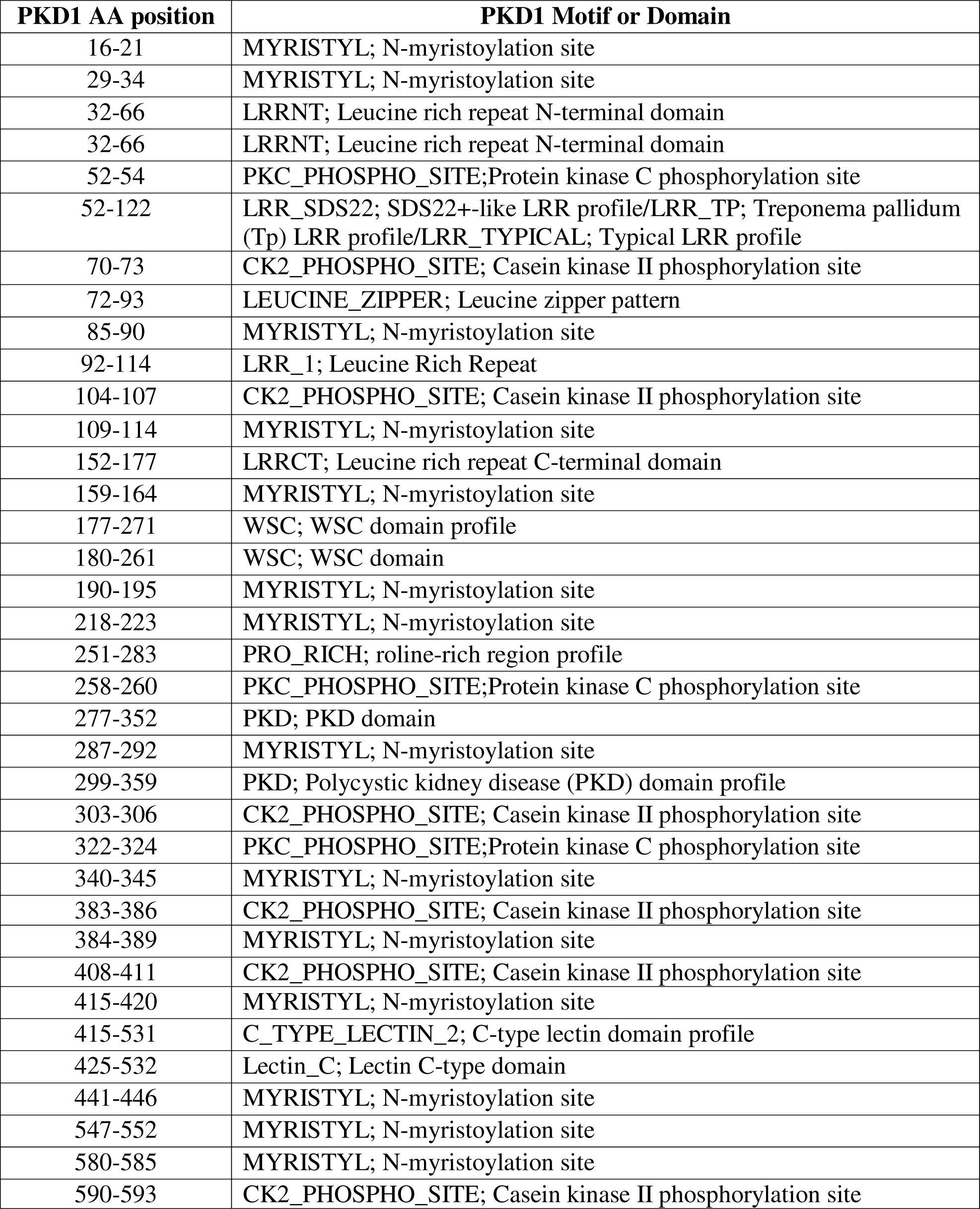
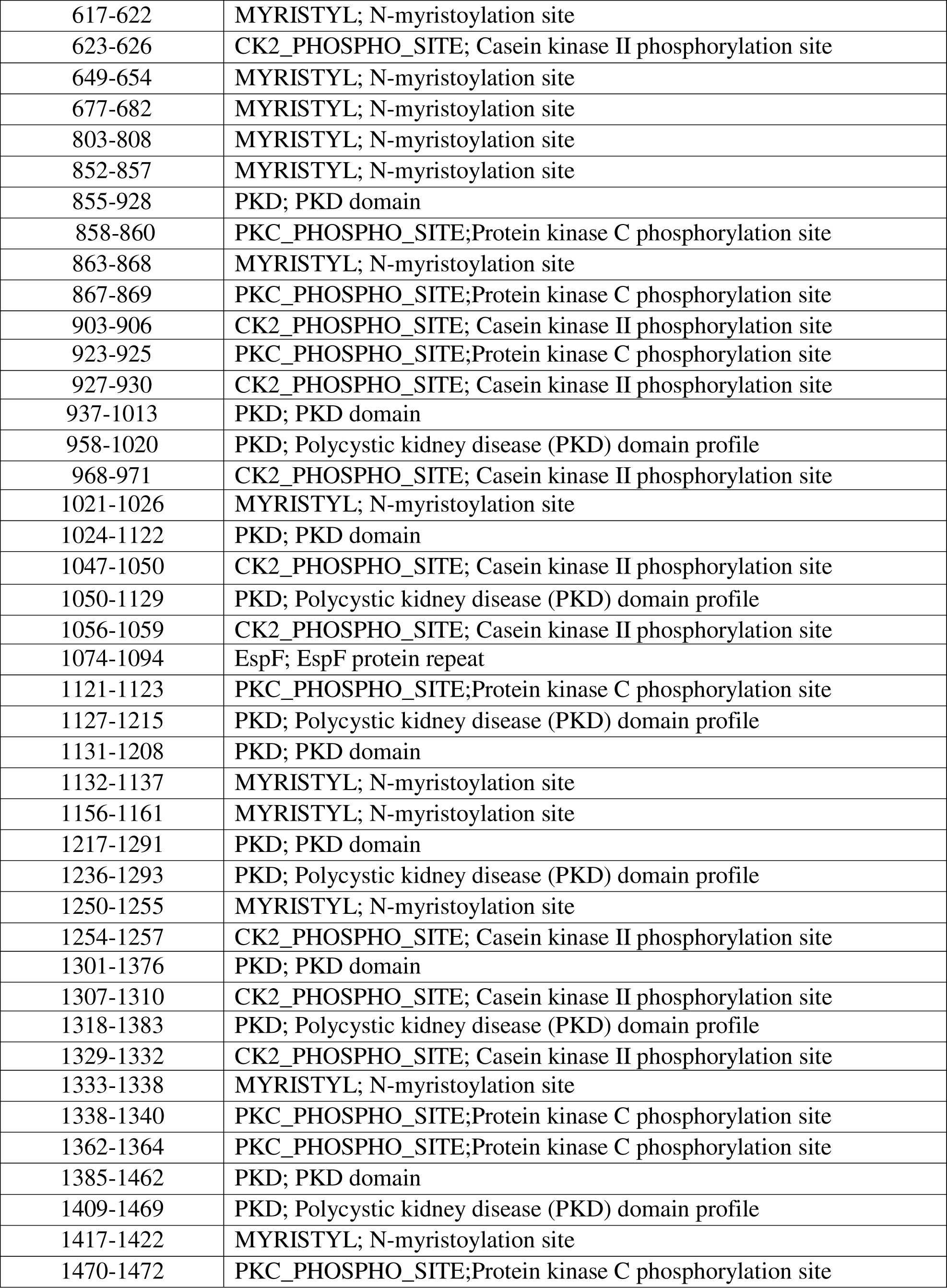
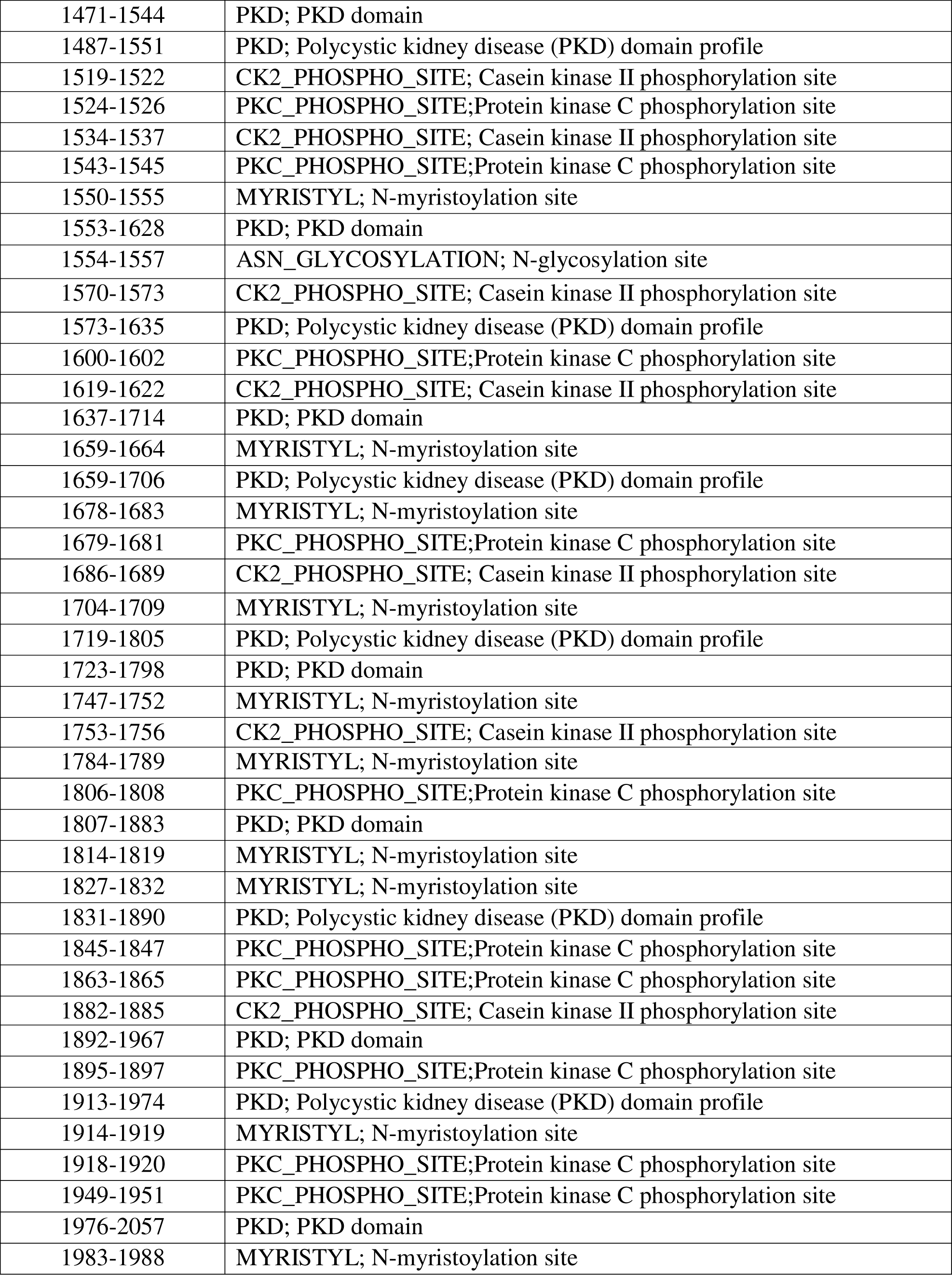
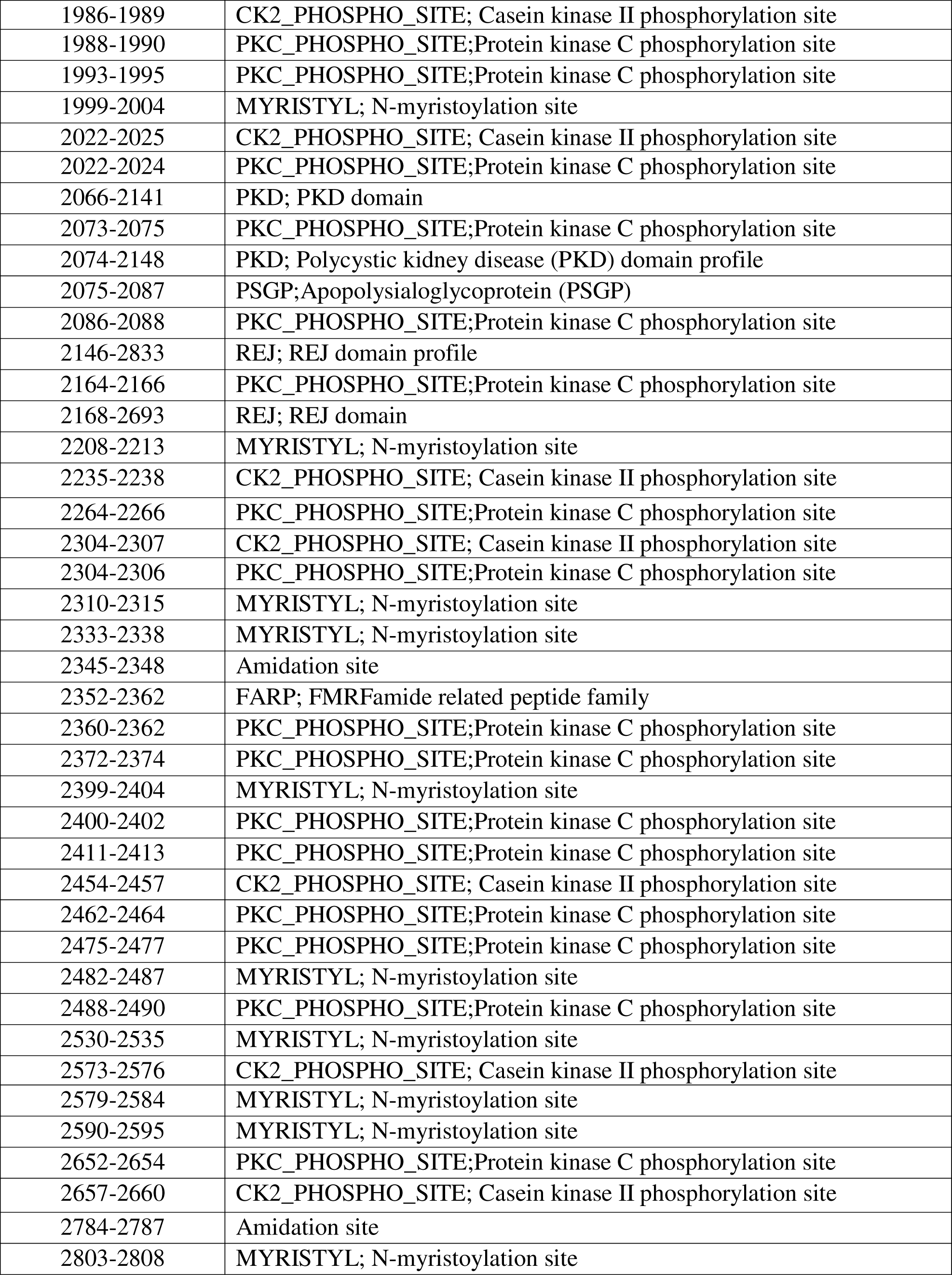
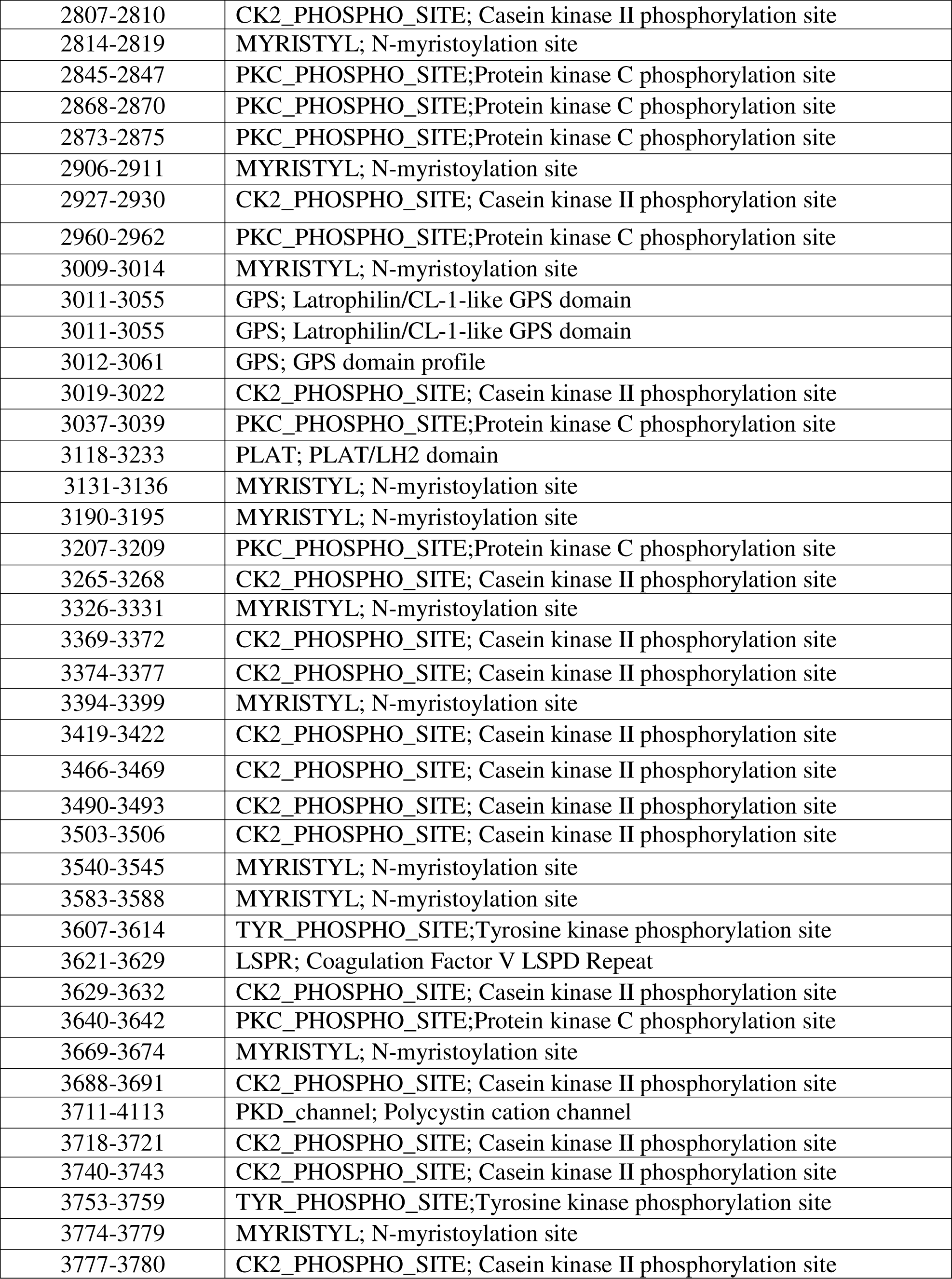
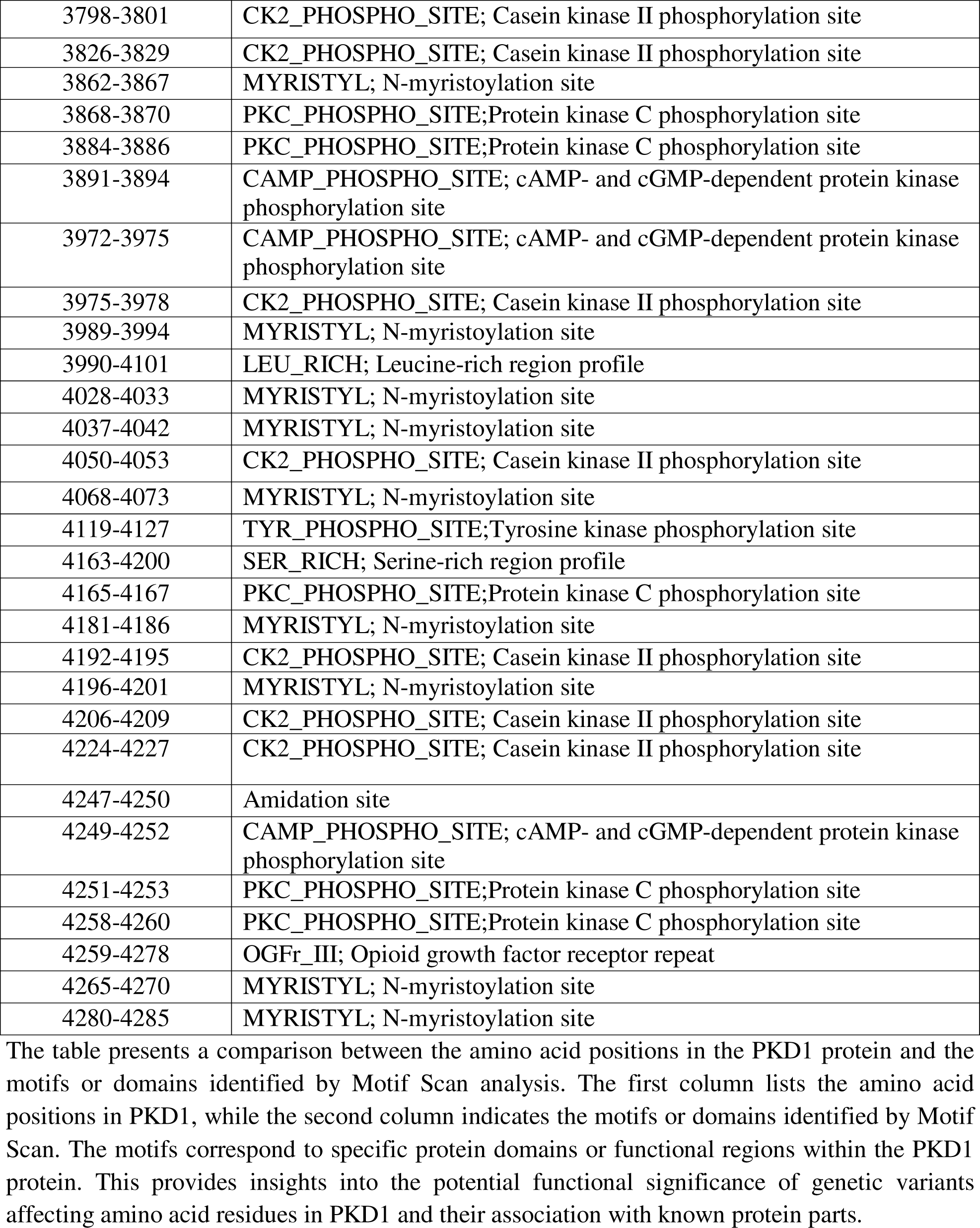
PKD1 Amino Acid Positions and corresponding Motifs or Domains.

### *PKD2* Gene Variants

The variants identified in the *PKD2* gene are Intronic, Splice Site Region c.2358+5G>A in Exon 12, c.*174G>T in 3’ UTR (Untranslated Region), Missense p.Ser370Asn (c.1109G>A) variant in Exon 5 (Table 2). The variants c.2358+5G>A and c.*174G>T with ACMG classifications and predicted to be deleterious and benign, respectively. These variants were reported in population databases and were categorized as uncertain significance in ClinVar.

### Predictions and Availability in Databases

Predictions of pathogenicity were based on ACMG criteria, MUT Assessor scores, and various computational tools (Table 3). Variants were cross-referenced with population databases such as Gnomad genome, ExAC and 1000G, as well as ClinVar, to assess their prevalence and significance. Some variants were not found in population databases and had limited evidence in literature or ClinVar (Table 3).

### WES of a Trio

*MIOX* was filtered out to have the frameshift-truncation deletion [c.32del/p.Leu11ArgfsTer61] shared by both affected mother and daughter carrying *PKD1* c.2899T>C, p.Trp967Arg missense variant. The heterozygous deletion variant of the *MIOX* was identified as the top possible disease associated variant. This variant was cross-validated bi-directionally by Sanger sequencing (figure 2A). The frameshift deletion leads to truncated protein of 70 amino acids as compared to wild protein of 285 amino acids. The alignment of 285aa long normal and 70aa long mutated MIOX protein sequence is shown in figure 2B. As per the information available on the databases (Proteomics DB and MOPED) the MIOX protein specifically expresses in the kidney (figure 3C).

**Figure 2:**
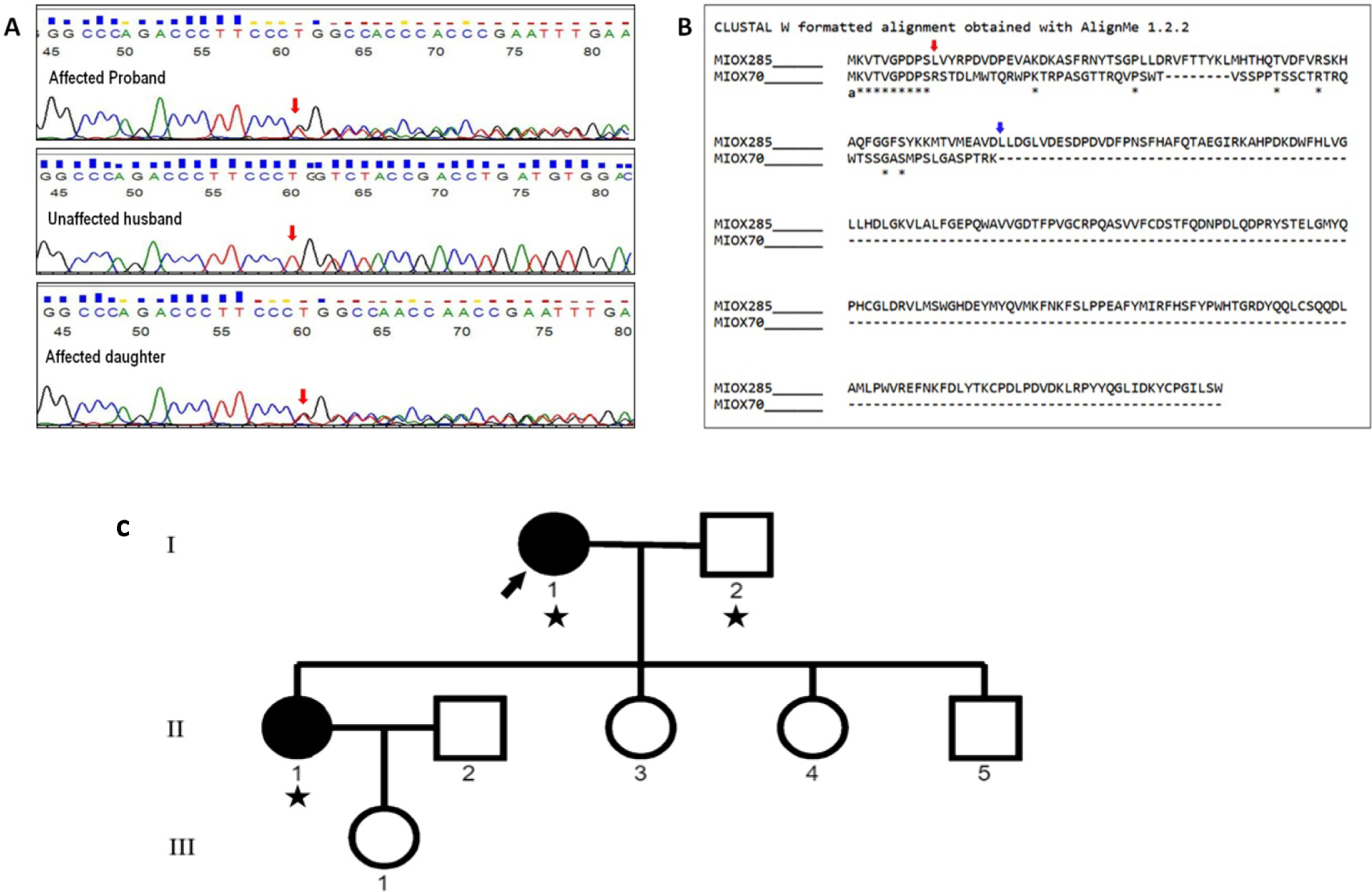
Sanger sequencing confirmation and alignment of normal (Miox285) and mutated (Miox70) protein sequence. A) Sanger sequencing confirmed the heterozygous deletion variant in the proband and affected daughter. Red arrow points the variant base. B) The alignment of normal (Miox285) and mutated (Miox70) protein sequence The Asterisks (*) represents the matched amino acids. Red and blue arrows indicate the amino acid change due to nucleotide change and truncation position respectively. Frame-shift deletion leads to truncated protein of 70 amino acids as compared to wild protein of 285 amino acids. C) Pedigree of the ADPKD subject. Star (★) indicates the trio undergone WES.

**Figure 3:**
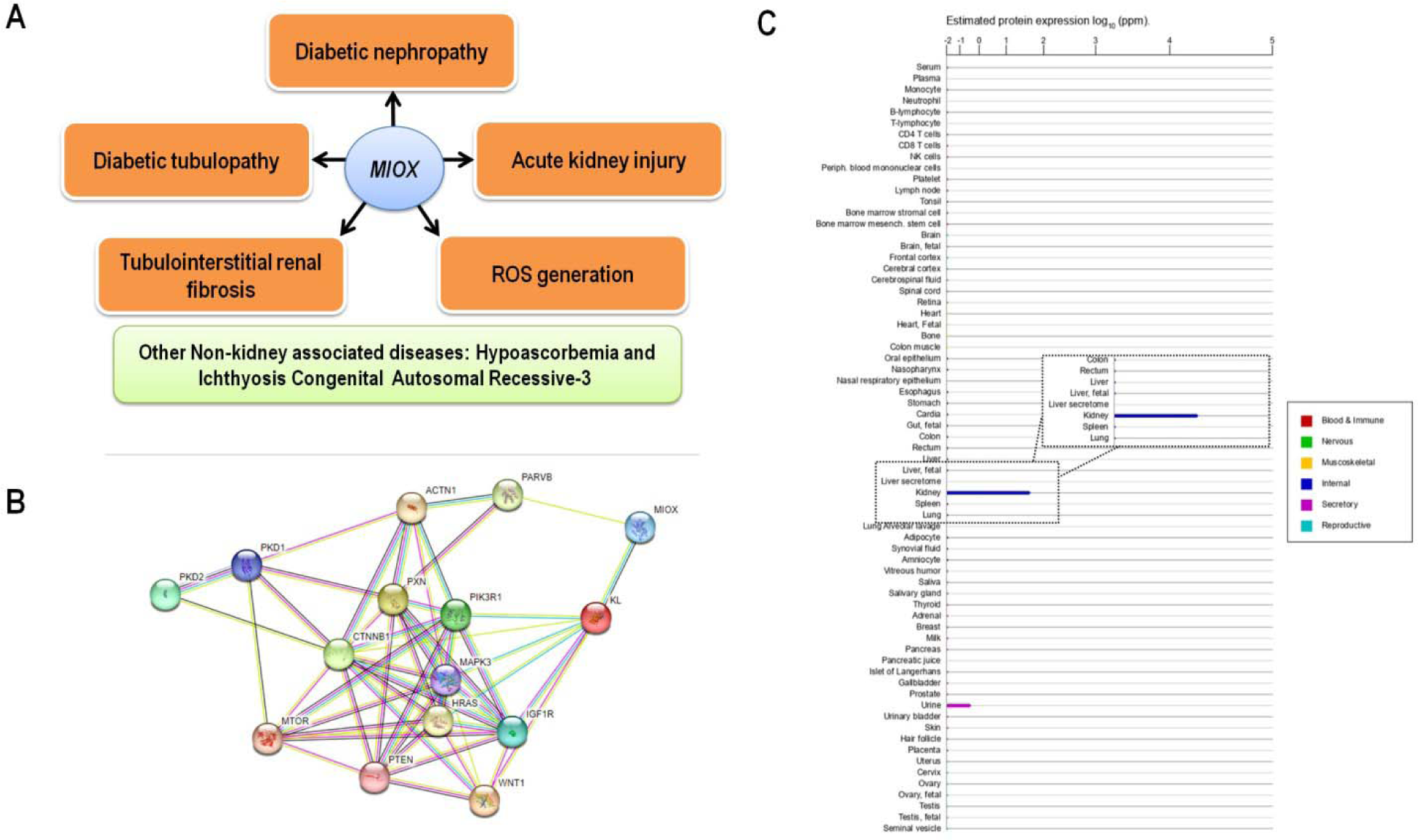
Role of MIOX protein. A) Association of *MIOX* in kidney and non-kidney diseases. B) Representative image depicting different types of interactions among common genes in *PKD1* and *MIOX* STRING interaction networks. C) Integrated Proteomics: protein expression in normal tissues and cell lines from Proteomics DB and MOPED for *MIOX*.

### Differential Gene Expression Analysis

The normalized gene expression data of the cysts is shown in figure 4A. The analysis revealed that the cyst samples assembled together as a distinct group, while the minimal cystic tissues and normal renal cortical samples sorted together as a separate group (figure 4B). This indicates the similarity in the gene expression pattern between renal cysts of varying sizes, as well as between minimal cystic tissues and normal renal cortical tissue. In addition, the expression profile analysis of *MIOX* reveal that its expression level is reduced in cystic samples compared to minimal cystic tissues and control tissue samples, suggesting a potential role of this gene in the development of polycystic kidney disease.

**Figure 4:**
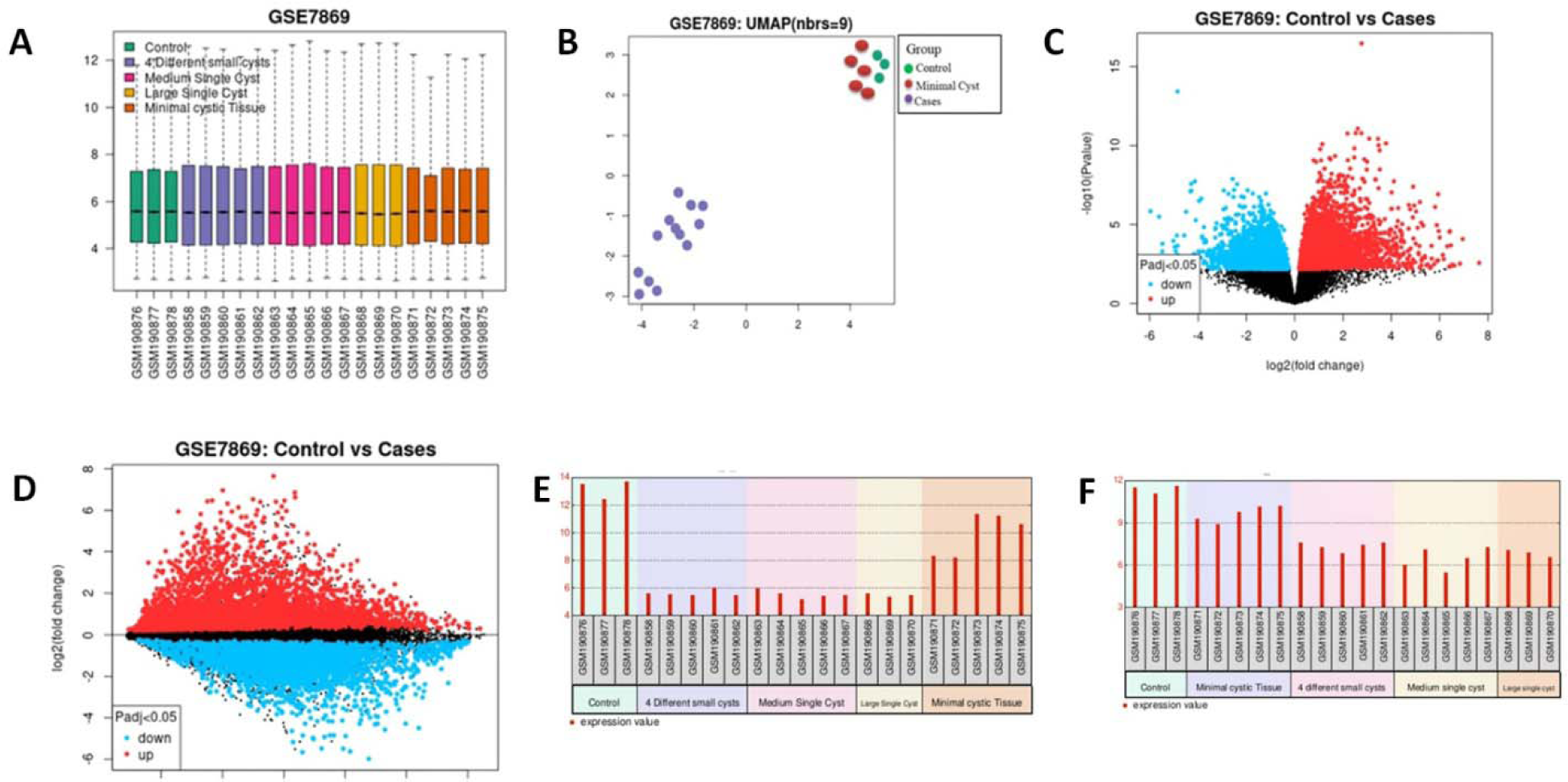
*MIOX* differential expression study in PKD1 cysts. A) Samples are colored based on groupings. Median-centered values indicate that the data are cross-comparable and normalized. B) The dimension reduction method known as uniform manifold approximation and projection (UMAP) representing the relationships between different cyst samples. C & D) The differentially expressed genes are depicted by volcano and mean difference plot. The red and sky-blue dots reflect up regulated and down regulated genes respectively. E) Expression of *MIOX* in cystic samples of ADPKD patients is down regulated as compared to minimal cystic tissues and control tissue samples. F) *KL* gene expression is down regulated as compared to minimal cystic tissues and control tissue samples.

## Discussion

ADPKD is a genetic disorder characterized by progressive cyst formation and enlargement of the kidneys, often leading to end-stage kidney disease. The genetic basis of ADPKD is heterogeneous, with PKD1 (∼75% of cases) and PKD2 (∼15% of cases) being the most common loci, and *GANAB, DNAJB11, ALG9,*and *IFT140* being minor genes [4, 5, 19]. PKD is a ciliopathy, a disease associated with defects in the function of primary cilia, and many syndromic ciliopathies exhibit a PKD phenotype [20]. *PKD1* pathogenic variants are associated with ADPKD, which manifests as renal and liver cysts, intracranial aneurysms, pain, nephrolithiasis, and end-stage renal disease. Understanding the genetic basis of ADPKD is crucial to understand its pathogenesis, identifying potential therapeutic targets, and improving clinical management strategies. Improving outcome, preventing complications, and initiating effective treatment rely heavily on accurate diagnosis. Molecular diagnosis also plays a critical role in the management of the diseases. WES is now becoming the choice of quick and accurate diagnosis for PKD and practicing personalized medicine. Given the complexity of the disease genetic testing in combination with clinical phenotypes offers definitive diagnosis contributing to better prognosis, clinical care, diagnosis in atypical cases, and family planning [12, 13, 21].

In this study, we conducted WES on a small cohort of 14 participants (from 12 families). Familial ADPKD was reported in all cases but one, indicating a clear hereditary pattern consistent with the autosomal dominant inheritance of the disease supporting the genetic basis of ADPKD. The imaging studies revealed multiple cysts of varying sizes in the kidneys. While bilateral involvement was predominant among the patients, one proband (individual coded 10_CD103) presented with unilateral kidney cysts, an uncommon finding in ADPKD [22]. Intriguingly, the sibling of this proband also exhibited unilateral kidney cysts at the teenage. ADPKD is typically known for bilateral involvement of the kidneys. This case may align with the unilateral renal cystic disease (URCD) which can be a manifestation of early-stage ADPKD particularly in pediatric cases [23]. In this case of unilateral ADPKD, the initial unilateral involvement emphasizes the need for careful diagnosis and regular long-term monitoring, as the disease may progress to asymmetric bilateral involvement over time. This supports the importance of considering ADPKD even in such cases ensuring appropriate management and follow-up. Liver cysts were noted in three cases. Patients reported a range of systemic symptoms associated with ADPKD. These included flank pain, which was a common complaint among the cohort, often attributed to cyst enlargement or hemorrhage within the kidneys. Generalized weakness, fatigue, kidney stone, appetite changes, nausea, anemia, hematuria, and other non-specific symptoms (Table 1) in the cohort reflect the systemic impact of the disease on overall health and vitality. Despite the presence of common features, the cohort displayed clinical heterogeneity in terms of age at presentation, symptomatology, and disease severity. This highlights the heterogeneity of ADPKD and the variability in disease presentation among affected individuals.

Analysis of the WES data identified several genetic variants within the *PKD1* and *PKD2* which are known to be major candidate genes of ADPKD. The variants in *PKD1* were more prevalent in our cohort, consistent with previous reports highlighting the predominant role of *PKD1* variants in ADPKD pathogenesis [17, 24]. Variants identified in *PKD1* included missense changes, nonsense variant, frameshift variants, and intronic splice site variants, each with varying degrees of pathogenicity as classified in ClinVar. The *PKD2* variants were also detected, albeit less frequently, emphasizing the contribution of both genes to ADPKD.

The variants identified span various regions of the *PKD1* and *PKD2*, potentially impacting protein structure and function. The table 1, 2 and 3 presents clinical and genetic information of patients with ADPKD, detailing the approximate age at diagnosis, gene, variant, effect of variant, exon number, and protein region affected and predictions using various tools. Individual 6_CD66 harbored a frameshift variant in the exon 29 of the *PKD1* gene (p.Ser3305fs*84, c.9914_9915delCT) which is present in between the LH2 and PKD cation channel region of PC1 protein. This patient, diagnosed at age early 50s, presented with bilateral ADPKD, characterized by multiple cysts and calculi in both kidneys (Table 1-3). Clinical symptoms included flank pain and blood in urine, indicating more severe disease manifestations. Another frameshift mutation was observed in individual 7_CD73, with a variant in *PKD1* exon 15 leading to a premature stop codon (p.His1347fs*83, c.4041_4042delCA). This will lead to a truncated protein upto PKD 7th domain. This patient diagnosed at late teenage, exhibited bilateral ADPKD and reported symptoms of flank pain, indicating early-onset. Both patients’ kidney sizes were larger than average, indicating a more advanced stage of the disease. Individual 8_CD78 (ADPKD with hepatomegaly), also diagnosed at late teenage, exhibited frameshift deletion in the exon 23 of *PKD1* gene impacting in REJ domain profile region. The presence of a frameshift variant in exon 15 and a larger deletion in exon 23 may have contributed to the early onset and severity of the disease phenotype. Individual 4_CD25 harbored a frameshift variant p.Gln149fs*141, c.445delC in *PKD1* exon 4 causing a frameshift and deletion in the region between the LRR1 and WSC domain. This patient diagnosed at early 40s, had bilateral ADPKD with rare occurrences of symptoms such as flank pain and blood in urine.

A heterozygous nonsense variant, c.1777G>T (p.Glu593*), in exon 9 of *PKD1* was identified in 11_CD99 diagnosed at early 30s. This variant introduces a premature stop codon, resulting in a truncated protein. The variant was absent in population databases, including gnomAD exomes and genomes however reported in publication (table 3).

Patient number 1_CD1 carried a missense variant in exon 24 of the *PKD1* gene (p.Glu2937Lys, c.8809G>A), classified as of uncertain significance, leading to a non-synonymous coding change in the REJ domain profile region, possibly affecting protein folding and function. This patient, diagnosed at mid 40s, presented with bilateral ADPKD, with a reported family history of the disease. Notably another patient 9_CD98, late 20s, exhibited grossly enlarged kidneys carried homozygous missense variant c.6928G>A (p.Gly2310Arg) in the *PKD1*, affecting the REJ Domain of PC1. Homozygosity for missense *PKD1*, as seen in this case, may contribute to the manifestation of early ADPKD, possibly through mechanisms involving gene dosage. Reports of homozygous mutations in *PKD1* and biallelic inheritance of missense variants highlight the role of gene dosage in determining disease severity [14, 25].

Patient 5_CD42 (diagnosed at late 20s) and patient 3_CD8 (diagnosed at late teenage) both carried intronic splice site variants in exon 21 of the *PKD1* gene (c.8017-3C>G) and exhibited bilateral ADPKD with multiple anechoic cysts. Patient 2_CD3, diagnosed at early 50s, experienced generalized body weakness, had synonymous and intronic variants in the *PKD2* gene, with reported bilateral ADPKD and multiple cortical and medullary cysts. The older age at diagnosis may suggest a slower disease progression and milder clinical phenotype, despite the presence of potentially pathogenic variants.

Liver cysts are a common extra-renal manifestation observed in patients with ADPKD. In our cohort, liver cysts were detected in three patients, notably in individual 8_CD78, who presented with multiple cysts in both kidneys, liver along with liver enlargement and simple cysts in right ovary. This patient exhibited symptoms of stomach ache, indicating potential compression effects of liver and kidney cysts. The presence of liver cysts in ADPKD patients can vary widely in terms of size, number, and associated symptoms [26]. While some patients may remain asymptomatic, others may experience discomfort or complications due to cyst growth and compression of adjacent structures.

The discrepancy in age at diagnosis between missense changes (diagnosis at mid 40s) and frameshift/nonsense change (diagnosis at late teenage and late 20s, respectively) within or near the REJ domain region suggests a potential correlation between mutation type and disease onset timing in ADPKD. The frameshift mutations in *PKD1* have been associated with more severe clinical presentations, including bilateral involvement and symptomatic flank pain. In contrast, patients with synonymous and missense variants tended to have milder disease manifestations with fewer reported symptoms [17, 24, 27, 28]. The variants identified in the *PKD1* affect various protein regions, each with its potential impact on the function of PC1 (Table 7). For instance, the non-synonymous and frameshift deletion variants in or near the REJ domain profile region possibly disrupt the signal transduction, which is crucial for cell-cell interactions and signal transduction through PC-1. The frameshift or non-synonymous variant in the the 3^rd^, 7^th^, 16^th^ PKD repeats may influence cellular processes mediated by these domains. Similarly, the frameshift variant between LRR1 and the WSC domain, the nonsense variant between the Lectin C-type domain and PKD 2^nd^ repeat could disrupt critical interactions involved in cell signaling and renal development [29]. The presence of the same intronic splice site variation in the *PKD1* gene was observed in two individuals within our cohort. Functional studies such as RNA sequencing or splice assays may provide insights into the impact of this intronic variation on *PKD1* mRNA splicing and its association with ADPKD pathogenesis. The identification of individuals carrying variations in both the *PKD1* and *PKD2* raises questions about potential synergistic effects or genetic modifiers influencing disease severity and progression as the patient is diagnosed at an early age (late-teenage). The exon number and affected domains appear to influence disease severity and diverse clinical phenotypes in ADPKD patients. However, further larger studies are needed to elucidate the specific functional consequences of variants in different exons and their impact on disease progression in ADPKD.

**Table 6:** Conserved Domains of PKD1 Protein.

| **Name** | **Accession** | **Amino acid Interval** | **E-value** |
| --- | --- | --- | --- |
| PCC super family (polycystin cation channel protein) | cl28216 | 97-2728 | 0.00E+00 |
| PKD channel super family (Polycystin cation channel) | cl37568 | 3713-4113 | 2.37E-106 |
| PLAT polycystin (PLAT/LH2 domain of polycystin-1 like proteins) | cd01752 | 3118-3237 | 8.86E-58 |
| GPS super family (GPCR proteolysis site, GPS, motif) | cl02559 | 3011-3060 | 3.08E-10 |
| PPP1R42 super family (protein phosphatase 1 regulatory subunit 42) | cl42388 | 54-127 | 6.08E-09 |

The table represents the conserved domains identified within the PKD1 protein using the NCBI Conserved Domains Database. Each domain is listed along with its corresponding accession number. These conserved domains provide information of the structural and functional characteristics of the PKD1 protein.

**Table 7:**
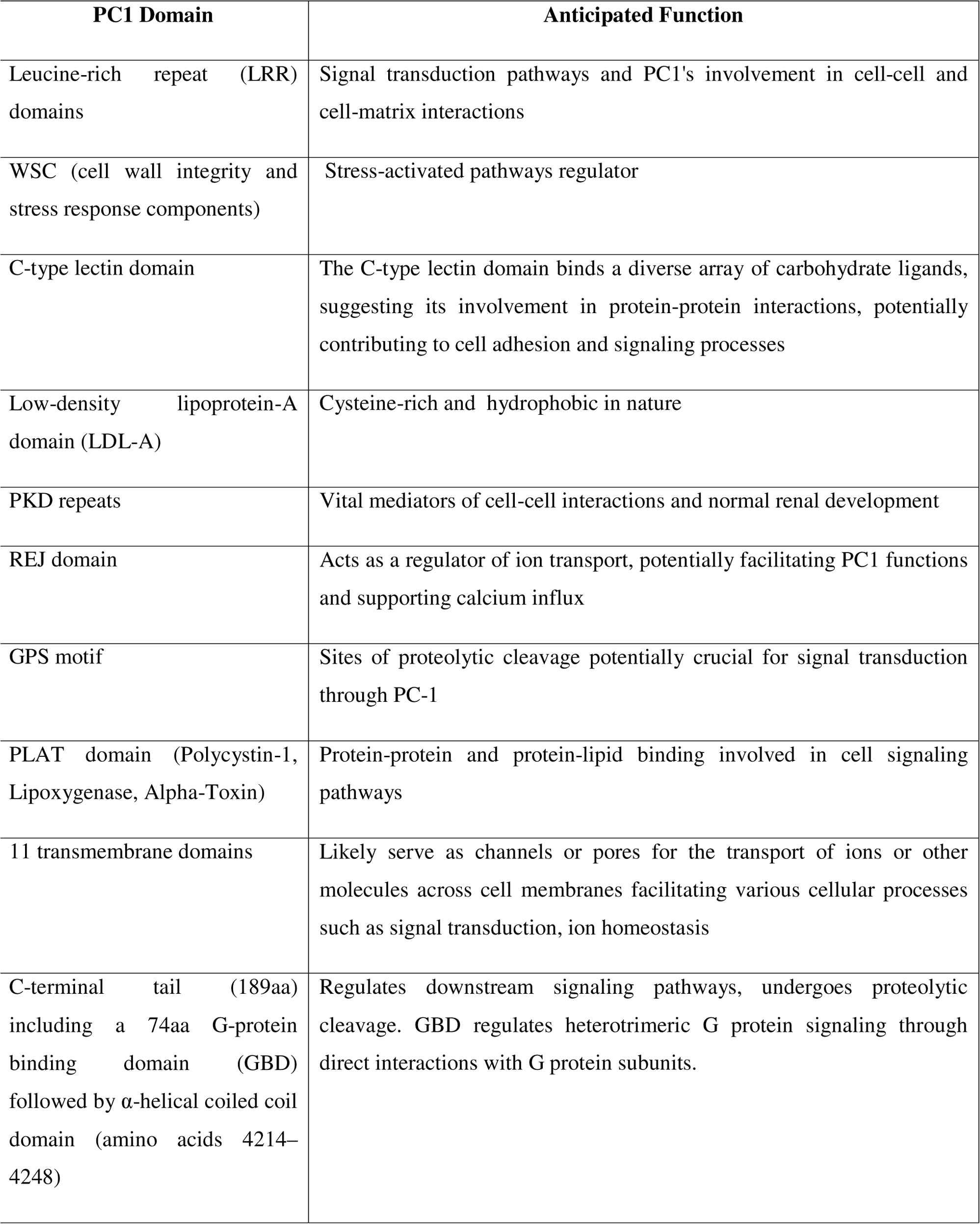
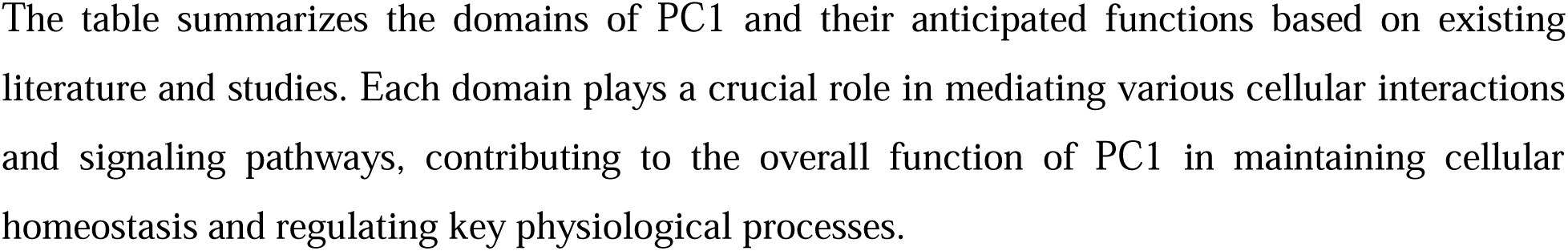
PC1 Domains and Anticipated Function.

In the other part of this study using WES of a trio (12_CD24 proband, CD24.1 proband’s husband, CD24.2 affected daughter) identified a missense *PKD1* variant shared by both affected mother and daughter. Interestingly, a heterozygous frameshift deletion [c.32del/p.Leu11ArgfsTer61, Exon 2] in *MIOX*, subsequently validated by Sanger sequencing, was also shared by both the affected persons. The variant was screened using the dominant inheritance pattern. This deletion has not been previously reported as an ADPKD related variation. The frameshift deletion leads to a truncated protein of 70 amino acids as compared to the wild protein of 285 amino acids which disrupts the binding site according to primary structure prediction. *MIOX* encodes for a 32Kda myo-inositol oxygenase (MIOX), which is a cytosolic enzyme expressed specifically in the renal proximal tubules, and is the first rate-limiting enzyme of myo-inositol catabolism. This enzyme is found to be upregulated in hyperglycemic conditions. MIOX is a single-domain protein with a mostly helical fold distantly related to the diverse HD domain superfamily. So far, studies have shown that MIOX plays a critical role in the metabolism of inositol, a carbohydrate molecule, and is involved in the regulation of glucose metabolism, insulin signaling, and oxidative stress response [16]. Studies have suggested the role of *MIOX* in insulin signaling in the renal proximal tubule cells and may play a role in the development of diabetic kidney disease [30]. Additionally, MIOX has been found to be involved in the regulation of oxidative stress, which can contribute to kidney injury in various disease states. *MIOX* promoter contains several response elements: oxidant-, antioxidant-, osmotic-, carbohydrate-, sterol-, response elements [30]. While the role of *MIOX* has been linked to acute kidney injury, tubulointerstitial renal fibrosis, diabetic-associated nephropathies, and reactive oxygen species (ROS) generation (figure 3A) [16, 31, 32], its role in ADPKD has not yet been reported. Recently, higher expression of *MIOX* has been reported in prostate adenocarcinoma [33].

As for *MIOX*, previous studies have explored the effects of overexpression and knockdown of *MIOX* in diabetic conditions. The current study identified a heterozygous frame shift deletion. This deletion leads to a premature termination of the MIOX protein, resulting in a truncated protein (70aa) that is much shorter than the wild-type MIOX protein (285aa). One possible impact of this deletion could be haploinsufficiency, where a single functional copy of the gene is insufficient to produce the required amount of functional protein necessary for normal cellular processes. This may lead to a reduction in the amount of functional MIOX protein below a threshold level, which could potentially trigger cyst formation. The other possibility could be a dominant-negative effect where the truncated protein created by heterozygous deletion may interfere with the function of the normal protein. This can disrupt the activity of protein complexes or signaling pathways, leading to disease.

The identification of a heterozygous frame-shift deletion in *MIOX* implies that this gene, along with the *PKD1* missense variant, may play a role in the early development of ADPKD in this specific trio, indicating a double heterozygous condition. The association of MIOX is further supported by the gene expression profile analysis of the data of cysts of variable sizes from GEO database, which shows that the expression level of *MIOX* gene is reduced remarkably in cystic samples compared to minimal cystic tissues (MCT) and control tissue samples (figure 4E). These results indicate that the down-regulation of *MIOX* expression may play a significant role in the development of PKD. In addition to this, the down regulation of Klotho (KL) protein in the reference study done by Song X et al. in 2009 [34] is noteworthy, as its direct STRING interaction with MIOX can be seen in our STRING interaction network (figure 3B & 4F). The *KL* encoded protein is a type-I membrane protein which is related to beta-glucosidases. Decreased expression of this KL protein has been reported in individuals with chronic renal failure. This suggests that the down-regulation of KL protein may be linked to the reduced expression of *MIOX* and further investigation into the relationship between these genes may shed light on their role in PKD pathogenesis. Overall, these findings highlight the importance of understanding the complex interplay between various genes and their expression profiles in the development and progression of PKD.

The all variants but one indentified in our cohort were different from those previously reported in our laboratory’s work [17]. The absence of identified variants in population databases and their submission in ClinVar without corresponding population frequency data highlight the need for population-specific genetic studies to fully capture the spectrum of disease-causing variants, particularly in understudied populations such as the Indian cohort in our study. Further functional studies, larger cohort analyses, and genotype-phenotype correlation studies would validate the pathogenicity of these variants and elucidate their impact on disease progression and clinical outcomes. The diversity of the variants suggests the genetic heterogeneity inherent in ADPKD, which may contribute to variability in disease presentation and progression among affected individuals. Understanding the genetic basis of ADPKD is crucial for improving diagnosis, prognosis, and the targeted therapies aimed at specific genetic mutations or pathways implicated in ADPKD pathogenesis which may benefit from the identification and characterization of novel variations, particularly those with potential functional significance.

### Study Limitations and future directions

The small sample size of our study cohort limits the generalizability of the findings to larger populations. Future studies with larger cohorts are needed to validate the significance of identified variations and elucidate their broader implications. Longitudinal studies incorporating clinical data and patient outcomes would establish genotype-phenotype correlations and assessing the prognostic value of identified variations in ADPKD progression.

## Conclusion

Our WES study expands the repertoire of genetic variations associated with ADPKD in the Indian population, providing insights into the genetic heterogeneity and complexity of this disease. Further, this study also identifies a frame-shift deletion [c.32del/p.Leu11ArgfsTer61] in *MIOX* besides *PKD1* missense variant, shared by both affected individuals in a trio which is not earlier reported in ADPKD. The frame-shift deletion results in a truncated protein with only 70 amino acids, which disrupts the binding site according to primary structure prediction. The differential gene expression analysis also showed reduced expression of *MIOX* in cysts samples. Further functional investigation is needed to determine the involvement of the *MIOX* variation in disease progression. The identification of this mutation in *MIOX* in an ADPKD trio study provides a potential avenue for further exploratory studies into the genetic underpinnings of ADPKD. These findings highlight the importance of understanding the complex interplay between various genes and their expression profiles in the development and progression of PKD. Further research at a larger scale is needed to elucidate the functional significance of identified variants, their contribution to disease pathogenesis, and their potential implications for clinical management and genetic counseling in ADPKD patients.

## Data Availability

All data produced in the present study are available upon reasonable request to the authors

## Acknowledgements

We acknowledge the funding support provided by the Department of Biotechnology (DBT) and the Senior Research Fellowship to first author obtained from the Indian Council of Medical Research (ICMR), India. We acknowledge Mr. Prashant Ranjan, ICMR-SRF, Centre for Genetic Disorders, Institute of Science, Banaras Hindu University, for his contribution in Differential Gene Expression analysis done in this study. We also extend our appreciation to the participants for their invaluable contribution to this study.

## References

1. Nobakht N, Hanna RM, Al-Baghdadi M, et al (2020) Advances in autosomal dominant polycystic kidney disease: a clinical review. Kidney Med 2:196–208

2. Torres VE, Bennett WM Autosomal dominant polycystic kidney disease (ADPKD) in adults: Epidemiology, clinical presentation, and diagnosis. Uptodate Waltham, MA Uptodate Inc http//www uptodate com (Accesed December 27, 2019)

3. Cornec-Le Gall E, Torres VE, Harris PC (2018) Genetic complexity of autosomal dominant polycystic kidney and liver diseases. J Am Soc Nephrol 29:13–23

4. Hopp K, Cornec-Le Gall E, Senum SR, et al (2020) Detection and characterization of mosaicism in autosomal dominant polycystic kidney disease. Kidney Int 97:370–382

5. Lanktree MB, Haghighi A, di Bari I, et al (2021) Insights into autosomal dominant polycystic kidney disease from genetic studies. Clin J Am Soc Nephrol 16:790–799

6. Lanktree MB, Guiard E, Li W, et al (2019) Intrafamilial variability of ADPKD. Kidney Int Reports 4:995–1003

7. Nishio S, Tsuchiya K, Nakatani S, et al (2021) A digest from evidence-based clinical practice guideline for polycystic kidney disease 2020. Clin Exp Nephrol 25:1292–1302

8. Mallawaarachchi AC, Lundie B, Hort Y, et al (2021) Genomic diagnostics in polycystic kidney disease: an assessment of real-world use of whole-genome sequencing. Eur J Hum Genet 29:760–770

9. Pei Y, Hwang Y-H, Conklin J, et al (2015) Imaging-based diagnosis of autosomal dominant polycystic kidney disease. J Am Soc Nephrol 26:746–753

10. Harris PC, Ward CJ, Peral B, Hughes J (1995) Polycystic kidney disease. 1: Identification and analysis of the primary defect. J Am Soc Nephrol 6:1125–1133

11. Cornec-Le Gall E, Olson RJ, Besse W, et al (2018) Monoallelic mutations to DNAJB11 cause atypical autosomal-dominant polycystic kidney disease. Am J Hum Genet 102:832–844

12. Du N, Dong D, Sun L, et al (2021) Identification of ACOT13 and PTGER2 as novel candidate genes of autosomal dominant polycystic kidney disease through whole exome sequencing. Eur J Med Res 26:1–8

13. Zacchia M, Blanco FDV, Trepiccione F, et al (2021) Nephroplex: a kidney-focused NGS panel highlights the challenges of PKD1 sequencing and identifies a founder BBS4 mutation. J Nephrol 34:1855–1874

14. Bergmann C, von Bothmer J, Brüchle NO, et al (2011) Mutations in multiple PKD genes may explain early and severe polycystic kidney disease. J Am Soc Nephrol JASN 22:2047

15. Burgmaier K, Kilian S, Bammens B, et al (2019) Clinical courses and complications of young adults with autosomal recessive polycystic kidney disease (ARPKD). Sci Rep 9:7919

16. Deng F, Sharma I, Dai Y, et al (2019) Myo-inositol oxygenase expression profile modulates pathogenic ferroptosis in the renal proximal tubule. J Clin Invest 129:5033–5049

17. Raj S, Singh RG, Das P (2020) Mutational screening of PKD1 and PKD2 in Indian ADPKD patients identified 95 genetic variants. Mutat Res Mol Mech Mutagen 821:111718

18. Ritchie ME, Phipson B, Wu DI, et al (2015) limma powers differential expression analyses for RNA-sequencing and microarray studies. Nucleic Acids Res 43:e47–e47

19. Senum SR, Li YSM, Benson KA, et al (2022) Monoallelic IFT140 pathogenic variants are an important cause of the autosomal dominant polycystic kidney-spectrum phenotype. Am J Hum Genet 109:136–156

20. McConnachie DJ, Stow JL, Mallett AJ (2021) Ciliopathies and the kidney: a review. Am J Kidney Dis 77:410–419

21. Odland D (2021) A patient perspective on genetic testing for ADPKD: the lack of complete genetic information, especially early in the course of the disease, is harming adult autosomal dominant polycystic kidney disease (ADPKD) patients. Clin. J. Am. Soc. Nephrol. 16:671–673

22. Shin C, Berliner L (2021) Case report: Atypical polycystic kidney disease. Radiol Case Reports 16:1643–1645

23. Gulzar M, Bakhsh M, Yonis H, et al Unilateral Renal Cystic Disease (URCD)

24. Pandita S, Ramachandran V, Balakrishnan P, et al (2019) Identification of PKD1 and PKD2 gene variants in a cohort of 125 Asian Indian patients of ADPKD. J Hum Genet 64:409–419

25. Audrézet M-P, Corbiere C, Lebbah S, et al (2016) Comprehensive PKD1 and PKD2 mutation analysis in prenatal autosomal dominant polycystic kidney disease. J Am Soc Nephrol 27:722–729

26. Kataoka H, Watanabe S, Sato M, et al (2021) Predicting liver cyst severity by mutations in patients with autosomal-dominant polycystic kidney disease. Hepatol Int 15:791–803

27. Singh S, Sreenidhi HC, Das P, Devi C (2023) Predicting the Risk of Progression in Indian ADPKD Cohort using PROPKD Score–A Single-Center Retrospective Study. Indian J Nephrol 33:195–201

28. Kataoka H, Fukuoka H, Makabe S, et al (2020) Prediction of renal prognosis in patients with autosomal dominant polycystic kidney disease using PKD1/PKD2 mutations. J Clin Med 9:146

29. Weston BS, Malhas AN, Price RG (2003) Structure–function relationships of the extracellular domain of the autosomal dominant polycystic kidney disease-associated protein, polycystin-1. FEBS Lett 538:8–13

30. Tominaga T, Dutta RK, Joladarashi D, et al (2016) Transcriptional and translational modulation of myo-inositol oxygenase (Miox) by fatty acids: implications in renal tubular injury induced in obesity and diabetes. J Biol Chem 291:1348–1367

31. Tominaga T, Sharma I, Fujita Y, et al (2019) Myo-inositol oxygenase accentuates renal tubular injury initiated by endoplasmic reticulum stress. Am J Physiol Physiol 316:F301–F315

32. Sharma I, Deng F, Liao Y, Kanwar YS (2020) Myo-inositol oxygenase (MIOX) overexpression drives the progression of renal tubulointerstitial injury in diabetes. Diabetes 69:1248–1263

33. Liu W, Xiang J, Wu X, et al (2022) Transcriptome Profiles Reveal a 12-Signature Metabolic Prediction Model and a Novel Role of Myo-Inositol Oxygenase in the Progression of Prostate Cancer. Front Oncol 12:

34. Song X, Di Giovanni V, He N, et al (2009) Systems biology of autosomal dominant polycystic kidney disease (ADPKD): computational identification of gene expression pathways and integrated regulatory networks. Hum Mol Genet 18:2328–2343

35. Kinoshita M, Higashihara E, Kawano H, et al (2016) Technical evaluation: identification of pathogenic mutations in PKD1 and PKD2 in patients with autosomal dominant polycystic kidney disease by next-generation sequencing and use of a comprehensive new classification system. PLoS One 11:e0166288

36. Rossetti S, Consugar MB, Chapman AB, et al (2007) Comprehensive molecular diagnostics in autosomal dominant polycystic kidney disease. J Am Soc Nephrol 18:2143–2160

